# Prevalence of Monogenic Aetiologies of Kidney Stone Disease in Selected and Unselected Kidney Stone Cohorts

**DOI:** 10.64898/2026.03.14.26348230

**Authors:** Catherine Lovegrove, Stefanie Croghan, Robert Geraghty, Holly Mabillard, Wael Asaad, Katherine Bull, Michael Holmes, Dominic Furniss, Alistair G. Rogers, John A. Sayer, Sarah A. Howles

## Abstract

**Background and objective:** Kidney stone disease (KSD) is common and distinct monogenic forms with well-defined treatment pathways exist. However, it is unclear which patients should undergo genetic testing and the significance of monoallelic variants in *SLC34A1, SLC34A3, CYP24A1, SLC7A9, and SLC3A1*. We aimed to define monogenic KSD inheritance patterns, determine the diagnostic yield of genetic testing in selected and unselected cohorts, and establish the utility of standard serum biochemistry in highlighting those at risk of monogenic KSD.

**Methods:** We compared genetic and phenotypic data from individuals with and without KSD in the UK Biobank to determine inheritance patterns. We established the “number needed to test” (NNTT) to identify monogenic KSD in the UK Biobank and a specialist adult kidney stone nephrology clinic where patients had genetic testing based on age, recurrence, and family history. We examined the utility of standard serum biochemistry in identifying monogenic KSD in both settings.

**Key findings and limitations:** Monogenic KSD associated with *SLC34A3, SLC7A9, and SLC3A1* is inherited in an autosomal dominant manner and monogenic KSD associated with *SLC34A1* and *CYP24A1* in an autosomal recessive manner. A monogenic diagnosis was made in ∼1% of KSD in the UK Biobank (NNTT∼90) and 15% of KSD in a specialist nephrology clinic (NNTT∼7). In both settings, standard serum biochemistry failed to identify individuals at risk of monogenic KSD.

**Conclusions and clinical implications:** Genetic testing for monogenic KSD has utility in specialist settings where there is clinical suspicion of an inherited disorder even in the absence of biochemical abnormalities.

## Introduction

Kidney stone disease (KSD) is a common condition that is associated with substantial morbidity, healthcare costs, environmental impact, and reduced quality of life^1–4^. Despite lifestyle modification, medical therapy, and surgical management, approximately half of affected individuals will experience recurrent disease ^2,5^.

KSD arises from a complex interplay of environmental exposures and genetic susceptibility, including a diverse set of Mendelian monogenic diseases ^6,7^. These monogenic disorders cause perturbations in calcium, phosphate, and vitamin D homeostasis (for instance infantile hypercalcaemia (IH), and hereditary hypophosphataemic rickets with hypercalciuria (HHRH)), abnormalities of oxalate metabolism (primary hyperoxaluria), cysteine transport (cystinuria), purine metabolism (including hereditary hyperuricosuria, hereditary xanthinuria, and adenine phosphoribosyltransferase deficiency), and acid-base handling (including distal renal tubular acidosis)^8^. Monogenic causes of KSD often present earlier in life, are frequently recurrent, and may be amendable to targeted therapies. Diagnosing these disorders has important implications for management, family screening, and genetic counselling^8–12^. However, identifying individuals with monogenic KSD is challenging. Standard biochemical evaluation, including serum calcium and phosphate measurement, lacks sensitivity, and variability in penetrance further complicates clinical recognition^13^. In the United Kingdom, genetic testing for KSD is performed using a panel-based approach (Nephrocalcinosis or Nephrolithiasis Panel R256, Genomics England) which includes genes with strong evidence for a Mendelian contribution to stone disease^9,14^. Despite the expert curation of this panel, uncertainty persists regarding the inheritance pattern of several monogenic forms of KSD^14^. This uncertainty complicates variant interpretation, clinical decision-making, and patient counselling.

Further uncertainty surrounds which patients should be offered genetic testing. Diagnostic yields for monogenic KSD vary by cohort, with diagnoses made in 3-7% of adult cases^15,16^ and up to 30% in those who form stones before 25 years of age^11^. Genetic testing for monogenic KSD is advocated by the European Association of Urology where there is clinical suspicion of an inherited or metabolic disorder^17^; however, the populations where this would be of value remain to be defined.

To address these uncertainties, we used clinical, biochemical and genetic data from the UK Biobank and a cohort of individuals with KSD attending a specialist nephrology clinic to clarify inheritance patterns of monogenic forms of KSD, assess the added diagnostic value of genetic testing over conventional biochemical analyses, estimate of the prevalence of monogenic forms of KSD, and define the number needed to test (NNTT) in selected and unselected cohorts.

## Methods

### Study cohorts

#### UK Biobank

The UK Biobank is a healthy volunteer study that includes health-related questionnaire data, physical measurements, biological samples, and linked medical records from approximately 502,000 participants^18^. We identified participants with KSD using diagnostic codes from the International Classification of Diseases (ICD) revisions 9 and 10, the Office of Population Censuses and Surveys Classification of Surgical Operations and Procedures (OPCS) revisions 3 and 4, Read versions 3 and 4, and self-reported codes (Supplementary Table S1). We excluded individuals whose genetic sex (data-field 22001) did not match their reported sex (data-field 31). We defined “recurrent” cases as those with a relevant KSD code occurring more than six months after the previous code.

#### Specialist kidney stone nephrology clinic

The Newcastle Upon Tyne NHS Foundation Trust specialist metabolic stone clinic reviews patients ≥ 18 years of age who have had KSD recurrence within 5 years of initial presentation, those who have had a KSD before 30 years or age, or individuals with an abnormality detected on 24-hour urine biochemistry following an agreed local referral pathway (Supplementary Figure S1). Patients were offered genetic testing if they were deemed to be at high risk of a monogenic form of KSD based on age, family history, and frequency of kidney stone recurrence.

### Genetic Testing and Variant Classification

#### UK Biobank

Exome sequencing and variant calling by the UK Biobank have been described^18,19^. The UK Biobank platform mapped whole-exome sequence (WES) reads to the hg38 reference genome using BWA MEM, generated per-sample gVCF files with WeCall, and aggregated them with GLnexus into a joint-genotyped, multi-sample project-level VCF (pVCF)^19^. The pipeline excluded SNPs with a read depth <7 and indels with a read depth <10. Only variants with ≥1 homozygous variant carrier or ≥ 1 heterozygous carrier with an allele balance of ≥ 0.15 for SNVs and ≥ 0.2 for indels were retained. Samples with discrepancies between genetic and reported sex, excess heterozygosity or contamination, inconsistencies between exome and array genotype calls, or ≥80% targeted bases achieving 20x coverage were omitted. Genetically identified sample duplicates and WES variants discordant with genotyping chips were also excluded.

Using Plink v2.0, we identified variants within the genomic regions of 35 genes associated with KSD, as listed in the NHS Genomic Medicine Service Nephrocalcinosis or Nephrolithiasis panel (Panel R256, Version 5.0, sign off date April 2025), based on Ensembl-defined coordinates, and annotated them using SnpEff^14,20–22^. We only included variants that met the following criteria: Minor allele frequency (MAF) ≤ 3%; genotype missingness < 5%; located in the canonical transcript; classified as “pathogenic” (P) or “likely pathogenic” (LP) according to ACMG guidelines, as annotated by InterVar^23,24^. Only *HNF4A* variants resulting in the R76W mutation were considered to have the potential to cause KSD as no other variants in this gene have been associated with kidney stones in the literature^25^.

#### Specialist kidney stone nephrology clinic

DNA samples were obtained from patients via a buccal salivary swab. Samples were processed at the Bristol Genetics Laboratory (national centre for gene testing for kidney stone patients), and tested against a 34 gene nephrocalcinosis or nephrolithiasis panel (R256, PanelApp Version 4.0, sign off date March 2023).^9^ Genes with high quality evidence supporting a causal monogenic role in KSD and their inheritance patterns are summarised in Supplementary Table S2. Genetic variants were classified by the Bristol Genetics Laboratory based on the American College of Medical Genetics and Genomics Criteria (ACMG) and those considered to be ‘pathogenic’ (P) or ‘likely pathogenic’ (LP) were considered to have the potential to cause disease.

### Definition of genetic inheritance modes

Most monogenic forms of KSD have well established monoallelic or biallelic inheritance models. However, the pathogenicity of monoallelic variants in *CYP24A1, SLC2A9, SLC34A1, SLC34A3, SLC3A1*, and *SLC7A9* is less certain (Supplementary Table S2)^14^. Thus, we sought to elucidate the inheritance mode of variants in these genes. We assessed genotype-phenotype correlations using KSD status, non-fasting serum phosphate, and albumin-adjusted serum calcium concentrations (*Adjusted calcium = Total calcium + 0.8 * (4.0 − Albumin))*. We considered associations in these genes under both monoallelic and biallelic models of inheritance. Under the monoallelic model, participants heterozygous for a P/LP variant in one of these six genes were classified as affected. Under the biallelic inheritance model, participants with two P/LP variants in the same gene were classified as affected. Compound heterozygosity was inferred if two variants were present in the same gene, as strand orientation could not be confirmed.

### Genotype-phenotype correlation in a specialist kidney stone nephrology clinic

All patients attending the Newcastle Upon Tyne NHS Foundation Trust specialist metabolic stone clinic eligible for ‘full screening’ (Supplemental Figure S1) were offered genetic, serum (calcium, urate, parathyroid hormone (PTH), bicarbonate, 25-hydroxyvitamin D, phosphate) and 24-hour urinary (calcium, urate, oxalate, citrate, phosphate, sodium, volume, pH +/- cystine if cystinuria suspected) biochemical testing. Genetic results were correlated with each patients’ KSD history, kidney stone analysis, and the findings of their biochemical testing. This was obtained from retrospective chart review of the Electronic Patient Record (EPR) system.

### Appraising the utility of genetic testing in kidney stone disease

To appraise the utility of genetic testing for KSD, we incorporated putative definitions for the inheritance modes for *CYP24A1, SLC2A9, SLC34A1, SLC34A3, SLC3A1*, and *SLC7A9* into the NHS Genomic Medicine Service Nephrocalcinosis or Nephrolithiasis panel (Panel R256) (Supplementary Table S2). We used the UK Biobank to represent an “unselected kidney stone cohort”. We calculated the number needed to test (NNTT) to identify a Mendelian diagnosis of KSD in this unselected cohort and in the Newcastle Upon Tyne NHS Foundation Trust specialist metabolic stone clinic cohort. To assess the value of genetic testing over standard serum biochemical analyses, we interpreted serum phosphate measurements according to the standard UK adult reference range of 0.8–1.5 mmol/L and albumin-adjusted serum calcium using a normal reference range of 2.20–2.60 mmol/L.

### Statistical analyses

Continuous variables were reported as medians with interquartile ranges (IQR) or means with standard deviation (SD). Categorical variables were reported as counts with percentages. Associations between categorical variables were analysed using Chi squared test, and associations between continuous variables were analysed using independent t-tests. All two-sided Chi-square tests were used to assess whether the prevalence of monogenic diagnoses differed between KSD cases (including both single and recurrent stone formers) and controls. Fisher’s exact test was used in instances where there were fewer than 5 counts. Statistical significance was defined as P<0.05, with values adjusted for multiple testing using the Bonferroni method. Statistical tests were performed in Rv4.1 and Rv4.2^26^. NNTT was calculated as (total number of individuals tested/number of people with genetic diagnosis). Multivariable logistic regression was used to determine predictors of monogenic diagnosis.

### Ethical approval

The specialist cohort was registered as an audit with Newcastle-upon-Tyne hospitals trust (Audit no.: 10337). Informed written consent for genetic testing was taken from participants. Genotyping was undertaken as part of the National Genomic Research Library study and approved by the East of England - Cambridge Central Research Ethics Committee (IRAS ID 278318 REC reference 20/EE/0035).

UK Biobank is approved by the National Information Governance Board for Health and Social Care and the National Health Service North West Centre for Research Ethics Committee (Ref: 11/NW/0382). This research was conducted using the UK Biobank Resource under application number 83942.

## Results

### Defining inheritance kidney stone disease inheritance patterns

The contribution of monoallelic variants in *CYP24A1, SLC2A9, SLC34A1, SLC34A3, SLC3A1,* and *SLC7A9* to the development of KSD requires clarification. To provide insight, we undertook genotype-phenotype analyses.

We identified 13,681 participants with KSD and 455,614 controls with WES data in the UK Biobank (Supplementary Table S3). Of these, 11,424 were recorded to have had a single stone event and 2,257 to have had recurrent disease. Individuals with KSD were more likely to be male, have higher serum concentrations of urate and creatinine, lower serum phosphate and 25-hydroxyvitamin D levels, and increased body mass index and waist-to-hip ratio (Supplementary Table S3). These metabolic and anthropometric trends were more pronounced amongst those with recurrent KSD compared to participants identified as having experienced a single episode of KSD. People with KSD had a marginally lower albumin-adjusted serum calcium concentrations than controls (2.281 mmol/L vs 2.276 mmol/L, P<0.001), however there was no difference in serum calcium concentrations between non-recurrent and recurrent stone formers (2.28 mmol/L and 2.28 mmol/L, respectively, P=0.15) (Supplementary Table S4).

We identified P/LP variants in *CYP24A1, SLC2A9, SLC34A1, SLC34A3, SLC3A1,* and *SLC7A9* as described above (Supplementary Table S5). Considering these genes, under a monoallelic model of inheritance only P/LP variants in *SLC34A3* were associated with KSD in the UK Biobank (Supplementary Table S6). Using a biallelic inheritance model, there was no association of P/LP variants in any of the six tested genes with KSD. This may be due to the low frequency of these variants in this population that was not enriched for rare disease. No P/LP variants in *SLC2A9* were identified in the UK Biobank, precluding further analyses of this gene.

*SLC34A1* and *SLC34A3* encode sodium-dependent phosphate transport proteins 2a and 2c, respectively, which are expressed in the proximal renal tubule. Biallelic mutations in *SLC34A1* cause infantile hypercalcaemia type 2 (IH2) and biallelic mutations in *SLC34A3* cause hereditary hypophosphatemic rickets with hypercalciuria (HHRH). IH2 is caused by increased renal phosphate excretion resulting in a reduction in serum FGF23 concentrations, activation of 1-α hydroxylase (an enzyme that activates 25-hydroxyvitamin D), and inhibition of 24-hydroxylase. HHRH is characterised by rickets, short stature, increased renal phosphate excretion, hypophosphatemia, hypercalciuria, increased gastrointestinal absorption of calcium and phosphate by similar mechanisms owing to an elevated serum concentration of 1,25-dihydroxyvitamin D, and suppressed parathyroid function^27–29^. To gain insight into whether monoallelic *SLC34A1* or *SLC34A3* P/LP variants affect calcium-phosphate homeostasis, we evaluated serum phosphate and calcium levels in UK Biobank participants.

Considering the whole UK Biobank cohort, carriers of SLC*34A1* P/LP variants (517 individuals) had a lower mean serum phosphate concentration than non-carriers (468,778 individuals) (1.14 mmol/L vs 1.16 mmol/L, P = 0.01, Supplementary Table S7). Individuals with KSD and carrying *SLC34A1* P/LP variants (15 individuals) also had a lower mean serum phosphate concentration than individuals with KSD without a P/LP *SLC34A1* variant (1.03 mmol/L vs 1.12 mmol/L, P = 0.03, Supplementary Table S8). There was no difference in albumin-adjusted serum calcium concentrations. Linear regression in the KSD cohort, adjusted for age, sex, serum creatinine, and albumin-adjusted serum calcium indicated a 0.11 mmol/L lower phosphate concentration in *SLC34A1* variant carriers compared to non-carriers (Supplementary Table S9).

Considering the UK Biobank as a whole, carriers of SLC*34A3* P/LP variants (853 individuals) had a lower mean serum phosphate concentration than non-carriers (468,442 individuals) (1.11 mmol/L vs 1.16 mmol/L, P= 7.83×10-18, Supplementary Table S6). Serum calcium concentrations were equivalent. Amongst 51 participants with KSD and a P/LP variant in *SLC34A3,* mean serum phosphate concentration was lower than individuals with KSD without a P/LP *SLC34A3* variant (1.06 mmol/L vs 1.12 mmol/L, P = 0.03, Supplementary Table S7). Linear regression in the KSD cohort, adjusted for age, sex, serum creatinine, and albumin-adjusted serum calcium indicated a 0.06 mmol/L lower phosphate concentration in *SLC34A3* variant carriers compared to non-carriers (Supplementary Table S10).

Biallelic mutations in *CYP24A1* cause IH type 1 (IH1). *CYP24A1* encodes the enzyme 24-hydroxylase, and pathogenic variants impair the inactivation of 1,25-dihydroxyvitamin D. This results in elevated circulating 1,25-dihydroxyvitamin D concentrations, increased intestinal and renal calcium absorption, and subsequent hypercalcaemia^30–32^. To assess whether monoallelic P/LP *CYP24A1* variants influence calcium homeostasis we evaluated calcium concentrations in UK Biobank participants.

Across the entire UK Biobank cohort, carriers of *CYP24A1* P/LP variants (n=906) had a slightly higher mean albumin-adjusted serum calcium compared with non-carriers (2.29 vs 2.28 mmol/L, P= 2.67×10-4, Supplementary Table S6). Among individuals with KSD, calcium concentrations were similar between carriers and non-carriers (P = 0.14; Supplementary Table S7). Linear regression in the KSD cohort indicated no difference in albumin-adjusted serum calcium concentrations in *CYP24A1* variant carriers compared to non-carriers (Supplementary Table S11).

The UK Biobank lacks specific phenotypes associated with *SLC7A9* and *SLC3A1*, which cause cystinuria types A and B, respectively, precluding further assessments of these genes^33^. However, we and others have previously reported that monoallelic variants in *SLC7A9* and *SLC3A1* are sufficient to cause cystine stones^33,34^.

Together, these findings indicate that monoallelic P/LP variants in *SLC34A3, SLC7A9,* and *SLC3A1* may cause monogenic forms of KSD with incomplete penetrance. However, there is no compelling evidence that monoallelic variants in *SLC34A1* or *CYP24A1* are sufficient to cause KSD. Therefore, for further our ongoing analyses we assumed an autosomal dominant inheritance model for KSD associated with P/LP variants in *SLC34A3, SLC7A9,* and *SLC3A1* and an autosomal recessive inheritance model for KSD associated with P/LP variants in *SLC34A1* and *CYP24A1* (Supplementary Table S2).

### Prevalence of monogenic disease associated with kidney stone disease in the UK Biobank

Across the 35 genes included in the NHS Genomic Medicine Service Panel R256 for nephrolithiasis/nephrocalcinosis, we identified 1,207 P/LP variants in 29 genes (730 pathogenic and 477 likely pathogenic); these were distributed across 29 genes (Supplementary Figure S2, Table 1).

**Table 1.**
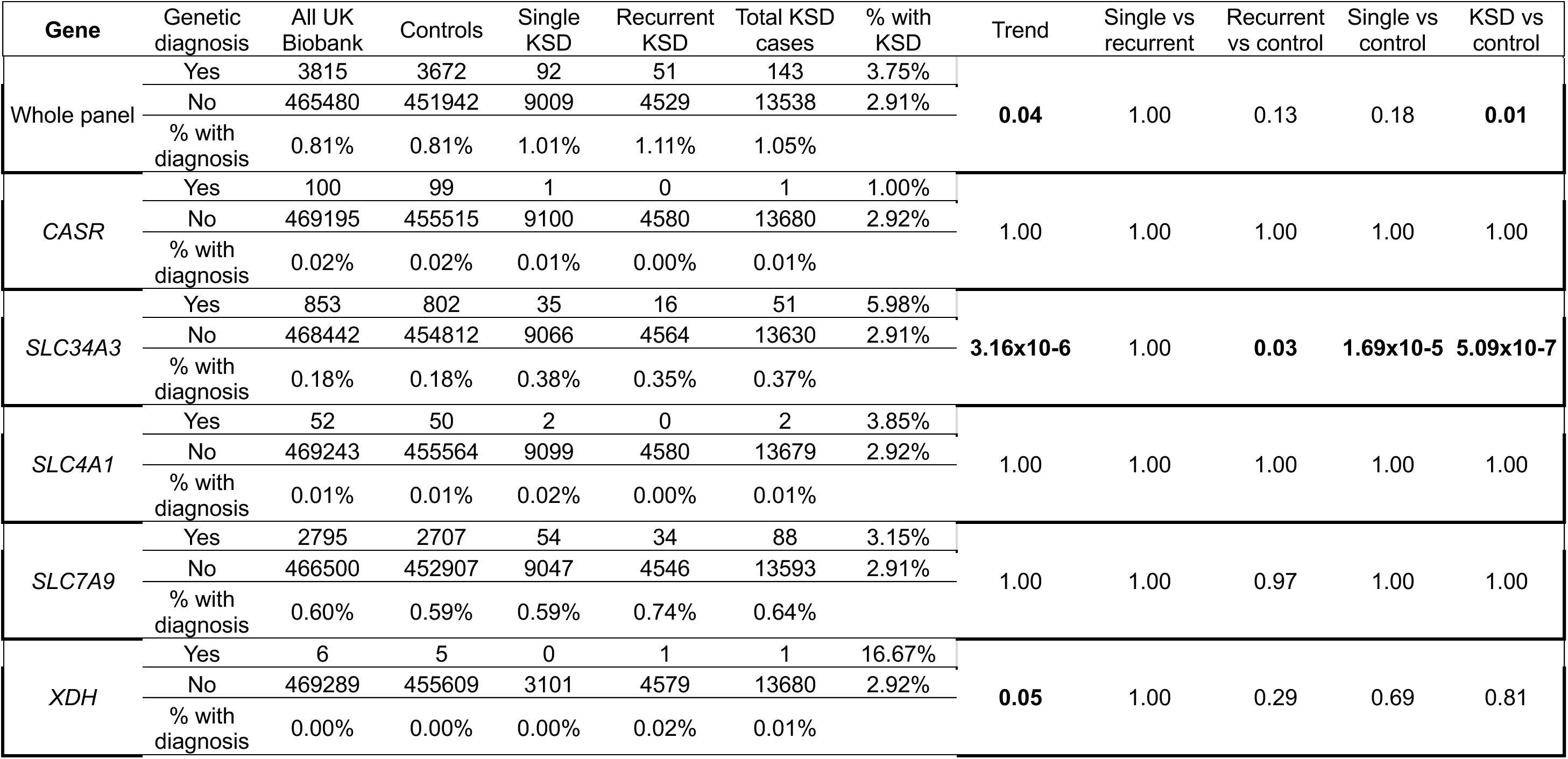
Proportion of Mendelian diagnoses of kidney stone disease in UK Biobank associated with the R256 Nephrocalcinosis or Nephrolithiasis panel.

Using the definitions described in Supplemental Table S2, there was a modest enrichment of Mendelian diagnoses in kidney stone cases compared to controls (P = 0.04; Table 1) with P/LP variants consistent with a Mendelian form of KSD identified in 143 (1.05%) KSD cases and 3,672 controls (0.81%) (Table 1). These variants were identified in *CASR, SLC34A3, SLC4A1, SLC7A9, and XDH* (Table 1, Supplemental Table S12).

The most common KSD Mendelian diagnosis was cystinuria secondary to variants in *SLC7A9,* (n=88, 0.64%), followed by monoallelic variants in *SLC34A3 (*n=51, 0.37%) leading to a *form fruste* of HHRH. P/LP variants consistent with Mendelian diagnoses of KSD were also present in the control group (*SLC34A3*: n=802, 0.18%; *SLC7A9*: n=2707, 0.59%) (Supplemental Table 13). One individual in the UK Biobank with KSD had a diagnosis of hereditary xanthinuria (*XDH*), two had a diagnosis of dRTA (*SLC4A1*), and one had disease associated with a Arg886Gln variant in the calcium sensing receptor (*CASR*). Loss- and gain-of-function heterozygous variants in *CASR* cause familial hypocalciuric hypercalcaemia type 1 (FHH1) and autosomal dominant hypocalcaemia type 1 (ADH1), respectively, both of which are associated with KSD. The Arg886Gln variant in *CASR* has not been functionally characterised, thus it is unclear whether this variant represents a potential diagnosis of FHH1 or ADH1 especially as the individual with this variant was normocalcaemic.

Among all putative compound heterozygotes, only one individual had KSD, this individual had two P/LP variants in *XDH*

In the UK Biobank, the age of individuals with a genetic finding consistent with a diagnosis of a monogenic form of KSD was comparable to those without a diagnosis (with, median age 61.0 years, interquartile range (IQR) 50.6-70.1; without, median age 61.4 years, IQR 50.0-69.7, Mann Witney U p=0.82).

### Prevalence of monogenic disease associated with kidney stone disease in specialist kidney stone clinic

Between May 2021 and Jan 2025, 243 patients attending the Newcastle Upon Tyne NHS Foundation Trust metabolic kidney stone clinic were screened for potentially disease-causing variants in genes included in the NHS Genomic Medicine Service Nephrocalcinosis or Nephrolithiasis panel. The mean age of the cohort was 45.5 ± 17.6 years, and the cohort comprised 124 men and 119 women (Table 2). A monogenic cause of KSD was identified in 37/243 (15%) patients (Table 3, Figure 1). Patients with a clinical diagnosis of cystinuria accounted for the most common molecular genetic diagnoses with P/LP variants found in *SLC7A9* in 16/37 cases with (9 biallelic and 7 monoallelic variants) and variants in *SLC3A1* seen in 8/37 cases (6 with biallelic and 2 with monoallelic variants). Among these 24 individuals, 18 had cystine stones confirmed on stone analysis. Elevated urinary cystine (via spot or 24-hour testing) was confirmed in 20 individuals. Of the 7 cases with heterozygous *SLC7A9* variants, 6 had evidence of cystinuria and 2 had cystine stones confirmed on stone analysis. Both patients with heterozygous *SLC3A1* variants had documented cystinuria (1871 and 2498 µmol/24 hours) and formed cystine stones, consistent with monoallelic variants in this gene being sufficient to cause cystine stones.

**Figure 1:**
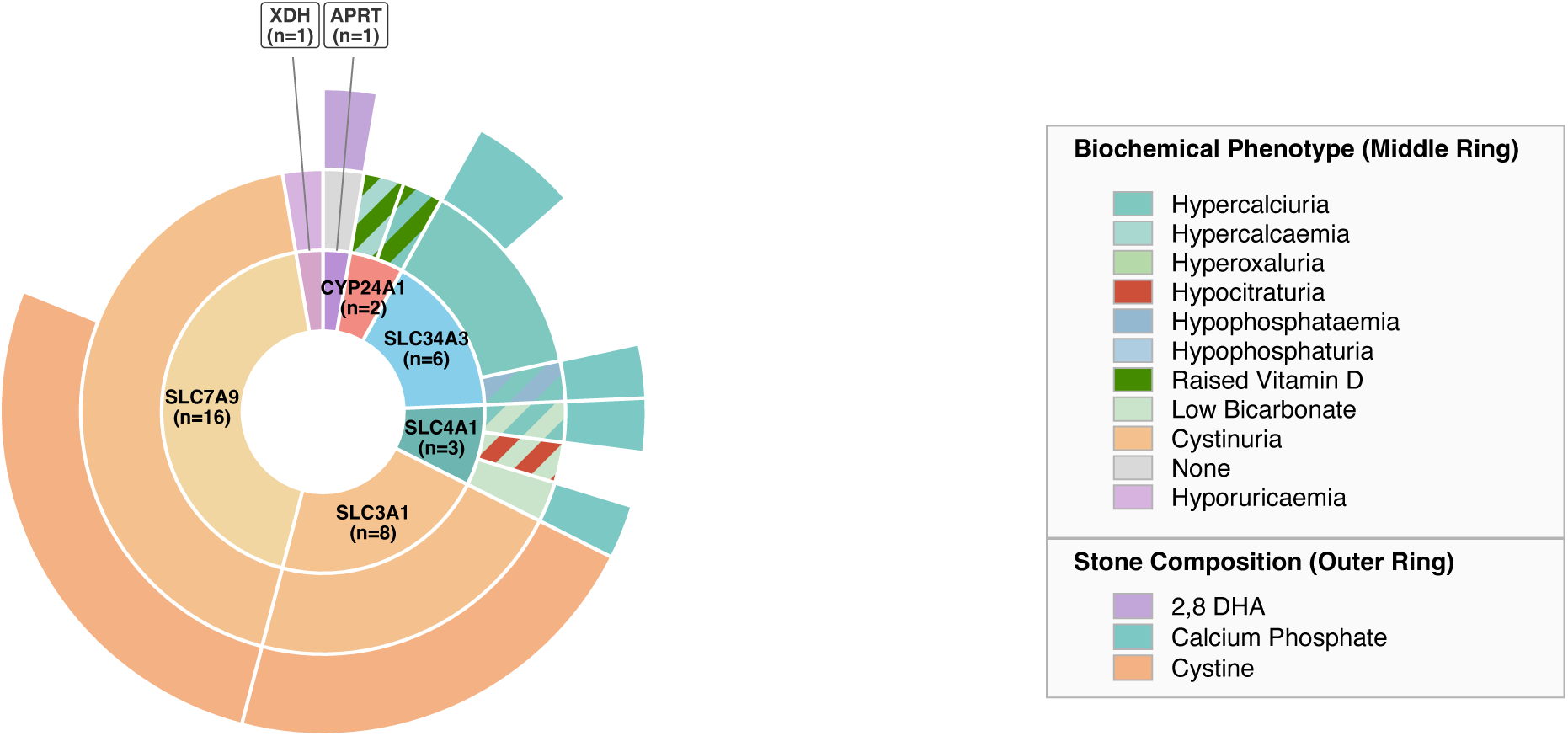
Distribution of genetic diagnoses and associated biochemical and stone phenotypes in the Newcastle Cohort.

**Table 2.** Demographics and monogenic diagnosis of Newcastle kidney stone cohort. Shown are baseline demographics and a multivariable logistic regression analysis of potential predictors of monogenic diagnosis.

|  | Monogenic Diagnosis<br>(P/LP variant) |  | OR (95% CI) | P |
| --- | --- | --- | --- | --- |
|  | Yes (n=37) | No (n=206) |  |  |
| **Female, n (%)** | 17 (45%) | 101 (49%) | 0.00 (-0.74 to 0.75) | >0.9 |
| **Age at First Stone event (Median, IQR)** | 29 (17-36) | 30 (21-46) | -0.03 (-0.06 to 0.00) | 0.03 |
| **Positive Family History, n (%)** | 13 (35%) | 137 (67%) | -0.16 (-0.97 to 0.65) | 0.7 |
| **Stone events in 5-years preceding testing, n (median, IQR)** | 2 (1-3) | 3 (2-5) | 0.01 (-0.97 to 0.65) | 0.7 |
| **Bilateral Stones, n (%)** | 9 (24%) | 61 (30%) | -0.07 (-0.89 to 0.76) | 0.9 |

**Table 3.**
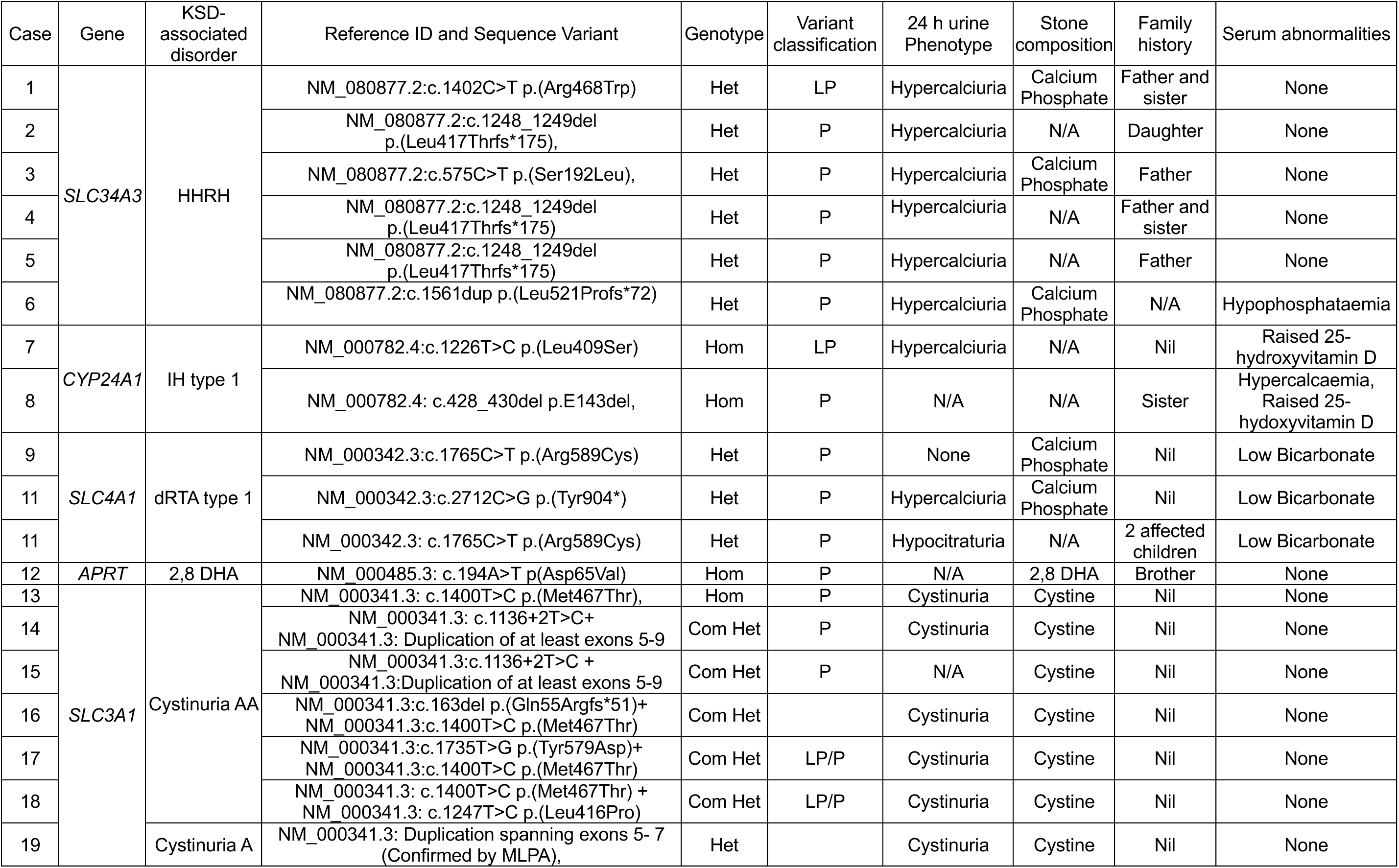
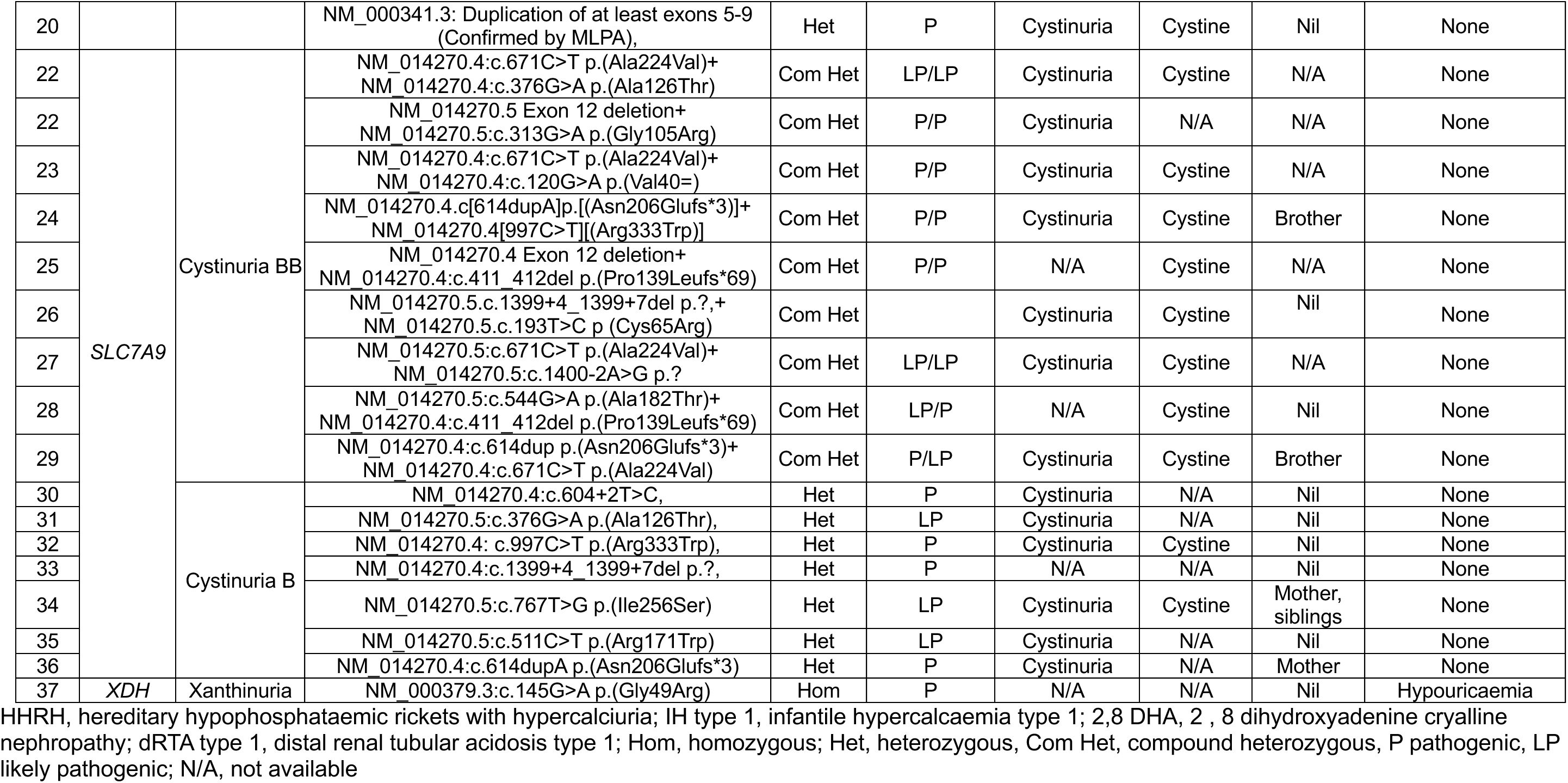
Pathogenic/likely pathogenic gene variants identified in a specialist kidney stone clinic cohort.

Six patients received a molecular diagnosis of a *form fruste* of HHRH and were heterozygous for P/LP variants in *SLC34A3*. Where stone composition was available, calcium phosphate stones were identified in 3/6 patients and hypercalciuria was confirmed in all 6 patients (Table 3). Two patients were diagnosed with IH1 with biallelic P/LP variants in *CYP24A1*, one of whom was hypercalcaemic with hypercalciuria and the other had a suppressed parathyroid hormone level, normal serum calcium but bilateral nephrocalcinosis and mild cystic kidney disease in keeping with known disease phenotypes. Both individuals had raised 25-hydroxyvitamin D concentrations. Three patients had monoallelic P/LP variants in *SLC4A1*, consistent with autosomal dominant distal renal tubular acidosis (dRTA). All three of these patients had a metabolic acidosis phenotype. Hypercalciuria or hypocitraturia was documented in 2 of the 3 cases, calcium phosphate stones were confirmed in 2 individuals, and nephrocalcinosis and renal cysts in the third (Table 3).

One patient with KSD had biallelic P/LP variants in *APRT*, conferring a diagnosis of APRT deficiency and another had biallelic *XDH* variants with a phenotype of recurrent kidney stones and severe hypouricaemia and an undetectable urine urate/creatinine ratio.

Multivariable logistic regression analyses indicated that younger age at first presentation may predict the presence of a monogenic KSD. However, there was no evidence to suggest that sex, family history, bilaterality of stones, or number of stone events predict monogenic KSD.

### Defining Clinical Utility of Genetic testing in unselected and selected kidney stone cohorts

To gain insight into the clinical utility of testing for monogenic forms of kidney stone disease in unselected and selected populations, we calculated the number needed to test (NNTT). In the UK Biobank, which may represent an unselected kidney stone clinic, the NNTT was 93 in those recorded to have had a single kidney stone episode and 90 in individuals with recurrent kidney stone disease (Table 4). P/LP variants in the Genomics England panel were predicted to account for < 0.25% of all KSD cases in the UK Biobank (Population Attributable Fraction, Supplemental Table S14). However, within a specialist metabolic kidney stone clinic where patients were selected for genetic profiling based on age, kidney stone recurrence, and family history, the NNTT was 7 (Table 4).

**Table 4.**
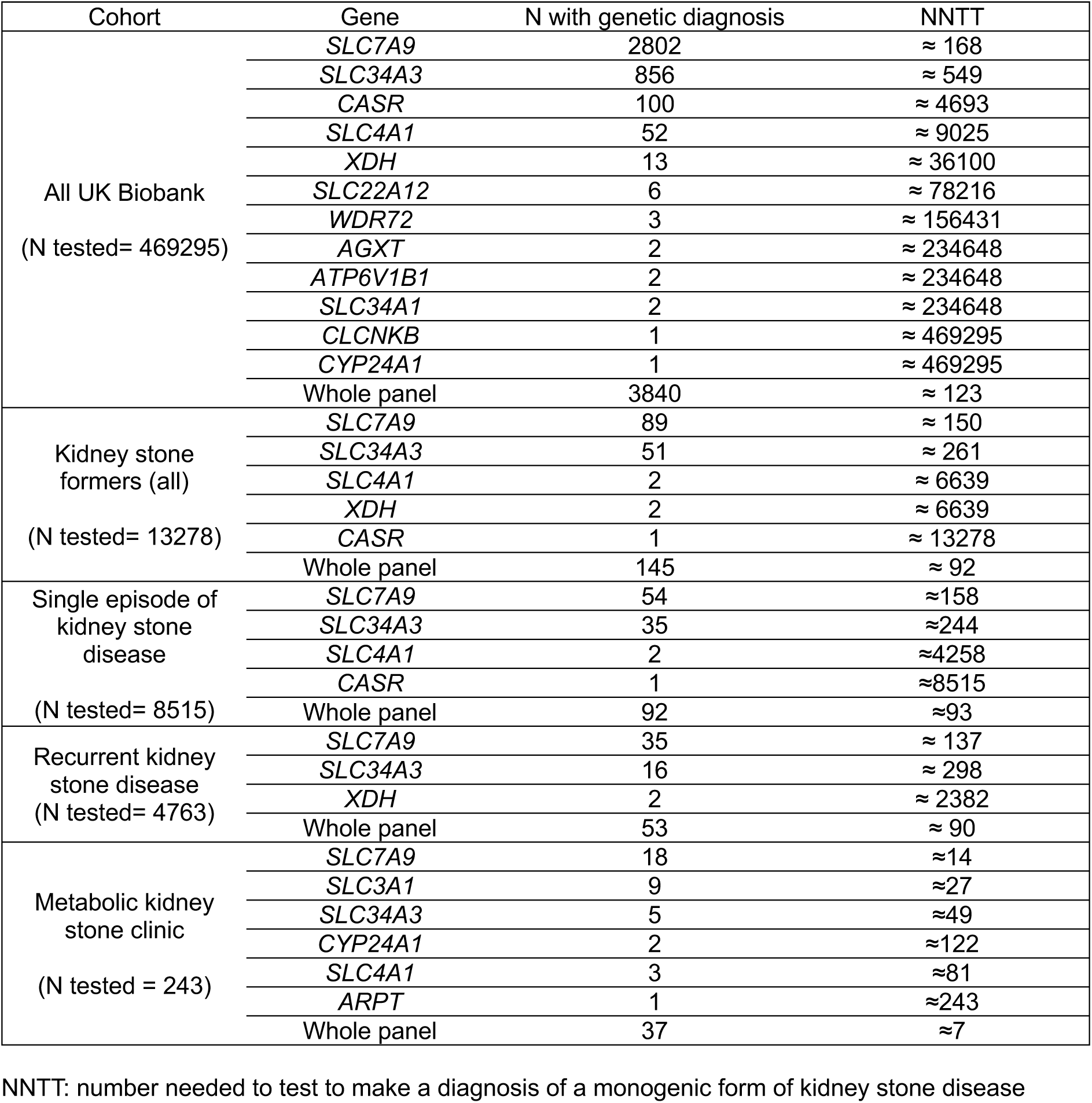
Number needed to test) to find a genetic diagnosis from the R256 Nephrocalcinosis or Nephrolithiasis panel.

### Biochemical profiling to detect monogenic forms of KSD

Both autosomal dominant and autosomal recessive forms of cystinuria and presumed autosomal dominant forms of HHRH were the most common monogenic forms of KSD in both UK Biobank and Newcastle cohorts. Individuals with cystinuria typically display elevated urinary cystine excretion and cystine stone formation. Thus, spot urinary cystine testing or stone analysis may have utility in screening for individuals for cystinuria.

Hypophosphataemia is often seen in association with HHRH secondary to variants in *SLC34A3*^27–29,35–39^. We hypothesised that simple biochemical profiling may highlight individuals at risk of HHRH to streamline diagnostic testing and evaluated phosphate levels in UK Biobank participants. Among 47/51 (92.12%) carriers of P/LP variants in *SLC34A3* with valid biochemistry data, 4/47 (8.51%) had hypophosphataemia (<0.80 mmol/L); 43/47 (91.48%) had phosphate levels in the normal range; none had elevated phosphate concentrations (Supplementary Table S15). Furthermore, hypophosphatemia was detected in only one individual who received a HHRH diagnosis in the Newcastle cohort (Table 3). These findings indicate that standard serum biochemistry would fail to highlight the majority of genetically confirmed *SLC34A3* cases.

## Discussion

In this study, we integrated population-scale genomic data with detailed phenotyping from a clinically enriched specialist cohort to define the clinical utility of genetic testing in KSD. Our findings provide three important insights. Firstly, we clarify the previously uncertain inheritance patterns of *SLC34A1, SLC34A3, CYP24A1, SLC7A9* and *SLC3A1.* Secondly, we demonstrate that monogenic forms of KSD are uncommon in unselected populations but substantially enriched in specialist KSD clinical settings. Third, we show that standard serum biochemical testing fails to identify individuals at increased risk of common monogenic forms of KSD, underscoring the added diagnostic value of genetic testing in selected high-risk populations.

Until now, a major barrier to effective use of genetic testing in KSD has been uncertainty concerning the inheritance patterns of several genes included in the diagnostic panel R256^14^. Using data from the UK Biobank, we show that monoallelic, P/LP variants in *SLC34A3* are associated with altered phosphate homeostasis and greater risk of KSD, supporting a clinically penetrant monoallelic disease model. These results align with previous studies reporting that P/LP heterozygous variants in *SLC34A3* contribute to hypercalciuria and nephrolithiasis, even in the absence of overt hypophosphataemia^28,39^.

In contrast, we found less evidence that monoallelic variants in *CYP24A1* or *SLC34A1* are sufficient to cause KSD. Although individuals heterozygous for *CYP24A1* and *SLC34A1* variants had marginally higher serum calcium and lower serum phosphate concentrations, respectively, overall differences were small and did not translate to an increased risk of KSD. These findings support monoallelic variants in *CYP24A1* and *SLC34A1* contributing to polygenic KSD inheritance and an autosomal recessive model for monogenic forms of *CYP24A1*- and *SLC34A1*-associated KSD. More detailed large-scale studies that include consideration of vitamin D metabolites including 1,25 dihydroxyvitamin D, which is not available in the UK Biobank, may provide additional insights.

While the UK Biobank lacked phenotypic information to clarify the inheritance pattern for *SLC3A1* and *SLC7A9* cystinuria-related KSD, our findings from specialist kidney stone nephrology clinic support previous literature reporting that monoallelic P/LP variants in both of these genes are sufficient to cause cystine stones, consistent with an autosomal dominant inheritance with variable penetrance^33^.

Together, our results suggest that inheritance follows an autosomal dominant model for *SLC34A3-, SLC3A1-,* and *SLC7A9-*associated KSD and an autosomal recessive model for *SLC34A1-* and *CYP24A1-*associated KSD, providing evidence to refine clinical genetic testing panels and improve variant interpretation and diagnostic accuracy.

In the unselected UK Biobank cohort, a monogenic KSD diagnoses could be made in ∼1% of individuals with KSD and there was only modest enrichment compared to controls consistent with incomplete penetrance. The population-attributable fraction was <0.25%, indicating that monogenic disease explains a minority of KSD cases at a population level, highlighting the limited clinical utility of genetic testing in unselected individuals with KSD.

In contrast, the diagnostic yield was far higher in a specialist nephrology kidney stone clinic, where genetic testing identified a monogenic cause of KSD in 37/243 (15%) patients, consistent with existing literature^11,15,39^. Whilst a younger age at presentation was statistically associated with findings consistent with a genetic diagnosis of KSD, median age at diagnosis was very similar (30 years vs 29 years). There was no evidence that other clinical features which have traditionally been used to select individuals for genetic testing including family history, bilaterality of disease, and number of stone events were able to predict the presence of monogenic disease, emphasising the limitations of phenotype-based selection alone. The disparity between diagnostic rates in unselected and selected cohorts translated into striking differences in testing efficiency. The NNTT exceeded 90 in unselected individuals with either single or recurrent stones but fell to 6 in the specialist clinic cohort. These findings support a targeted testing strategy, where genetic testing is reserved for patients with a clinical picture suggestive of a monogenic disorder in a specialist setting.

Our data demonstrates that standard serum biochemical screening alone will fail to identify the majority of genetically defined KSD cases. The most common genetic diagnosis was cystinuria in both selected and unselected cohorts. In the specialist clinic, this genetic diagnosis was supported by cystinuria and/or cystine stones in the majority of cases. Cystinuria does not cause alterations in blood biochemistry, is associated with chronic kidney disease, and is amendable to medical treatment thus identifying and treating this disorder has important implications.

Our data indicates that heterozygous variants in *SLC34A3* lead to a form fruste of HHRH and this was also a common monogenic KSD diagnosis in our cohorts. In the UK Biobank, fewer than 10% of individuals with a P/LP *SLC34A3* variant had overt hypophosphataemia, and only 1 individual who received an *SLC34A3*-associated HHRH diagnosis in the specialist clinic had detectable serum abnormalities of calcium-phosphate homeostasis. Several individuals with *SLC34A3* variants in the specialist clinic cohort were found to have hypercalciuria on 24-hour urine biochemistry, however this abnormality is detected in up to 50% of all individuals with KSD^40^ and therefore is not discriminatory in a clinical setting. These findings reflect the variable penetrance of biochemical abnormalities, highlight the limitations of single-time-point measurements, and indicate that genetic testing may reveal a monogenic form of KSD even when routine biochemical investigations are normal.

We note that the UK Biobank cohort is a healthy volunteer study and may not adequately represent the unselected KSD clinic which we sought to emulate in this study^18^. Participants in the UK Biobank are 40 to 69 years old, hence age-based stratification of NNTT was not possible in this setting. Furthermore, the lack of phenotypic data including urinary cystine excretion and stone composition precluded consideration of these phenotypes in this study. Finally, the Newcastle specialist clinic reflects referral practices at a single centre, and thus the results may not be generalisable.

Identifying monogenic forms of KSD may directly inform patient management; for example, *SLC34A3*-associated KSD may be successfully treated with oral phosphate supplementation. Furthermore, genetic diagnoses enable cascade testing and counselling of at-risk relatives^41^. Early identification of affected family members allows risk stratification and implementation of preventative strategies for KSD including increased fluid intake and dietary sodium restriction along with precision approaches based on genetic findings^42–44^.

Our results support genetic testing for monogenic forms of KSD in specialist settings where age, multiplicity of stones, recurrence, and family history raise suspicion of an inherited disorder. These data can be used to inform national genomic testing strategies and panel interpretation guidelines to reduce overcalling of variants and improve consistency across laboratories.

## Conclusions

Monogenic forms of KSD account for a small proportion of disease at a population level but are substantially enriched in specialist clinical settings. Monoallelic pathogenic variants in *SLC34A3*, *SLC3A1*, and *SLC7A9* confer clinically meaningful disease risk, whereas *CYP24A1*- and *SLC34A1*-associated KSD likely follow a biallelic inheritance model. Genetic testing provides diagnostic value beyond standard biochemical assessment, but its efficiency is dependent on patient selection. These findings support targeted genetic testing strategies and refined inheritance definitions to optimise the clinical management of KSD.

## Data Availability

All data produced in the present study are available upon reasonable request to the authors

## Rights retention statement

For the purpose of Open Access, the author has applied a CC BY public copyright licence to any Author Accepted Manuscript (AAM) version arising from this submission.

