## Supplementary material for "Prevalence of Monogenic Aetiologies of Kidney Stone Disease in Selected and Unselected Kidney Stone Cohorts": Supplmentary Appendix

**Supplementary tables (S1-S15)**

*Supplementary Table S1. Inclusion criteria for kidney stone disease association analyses in the UK Biobank.*

| Coding | Code | Definition |
|---|---|---|
| International Classification of Diseases (ICD) | ICD9- 7880 | Renal colic |
| | ICD9- 5920 | Calculus of kidney |
| | ICD9- 5920A | Calculus of kidney |
| | ICD9- 5921 | Calculus of ureter |
| | ICD9- 5929 | Urinary calculus, unspecified |
| | ICD10-N20.0 | Calculus of kidney |
| | ICD10-N20.1 | Calculus of ureter |
| | ICD10-N20.2 | Calculus of kidney with calculus of ureter |
| | ICD10-N20.9 | Urinary calculus, unspecified |
| | ICD10-N23 | Unspecified renal colic |
| Classification of Interventions and Procedures (OPCS) | OPCS3- 563.1 | Removal of renal calculus : nephrolithotomy |
| | OPCS3- 563.2 | Removal of renal calculus : pyelolithotomy |
| | OPCS3- 563.3 | Removal of renal calculus : removal without incision |
| | OPCS3- 580 | Ureterolithotomy |
| | OPCS3- 587.2 | Cystoscopic operation on ureter : lithotomy |
| | OPCS4- M06.1 | Open removal of calculus from kidney |
| | OPCS4- M07.1 | Ureteroscopic laser fragmentation of calculus of kidney |
| | OPCS4- M07.2 | Ureteroscopic extraction of calculus of kidney NEC |
| | OPCS4- M09 | Therapeutic endoscopic operations on calculus of kidney |

| | | |
|---|---|---|
| | OPCS4-M09.1 | Endoscopic ultrasound fragmentation of calculus of kidney |
| | OPCS4-M09.2 | Endoscopic electrohydraulic shockwave fragmentation of calculus of kidney |
| | OPCS4-M09.3 | Endoscopic laser fragmentation of calculus of kidney |
| | OPCS4-M09.4 | Endoscopic extraction of calculus of kidney NEC |
| | OPCS4-M09.8 | Other specified |
| | OPCS4-M09.9 | Unspecified |
| | OPCS4- M14 | Extracorporeal fragmentation of calculus of kidney |
| | OPCS4-M14.1 | Extracorporeal shock wave lithotripsy of calculus of kidney |
| | OPCS4-M14.8 | Other specified |
| | OPCS4-M14.9 | Unspecified |
| | OPCS4-M16·4 | Percutaneous nephrolithotomy |
| | OPCS4-M26.1 | Nephroscopic laser fragmentation of calculus of ureter |
| | OPCS4-M26.2 | Nephroscopic fragmentation of calculus of ureter NEC |
| | OPCS4-M26.3 | Nephroscopic extraction of calculus of ureter |
| | OPCS4-M27.1 | Ureteroscopic laser fragmentation of calculus of ureter |
| | OPCS4-M27.2 | Ureteroscopic fragmentation of calculus of ureter NEC |
| | OPCS4-M27.3 | Ureteroscopic extraction of calculus of ureter |
| | OPCS4- M28 | Other endoscopic removal of calculus from ureter |
| | OPCS4-M28·1 | Endoscopic laser fragmentation of calculus of ureter |
| | OPCS4-M28·2 | Endoscopic fragmentation of calculus of ureter |
| | OPCS4-M28·3 | Endoscopic extraction of calculus of ureter |
| | OPCS4-M28.4 | Endoscopic catheter drainage of calculus of ureter |

| | | |
|---|---|---|
| | OPCS4-M28.5 | Endoscopic drainage of calculus of ureter by dilation of ureter |
| | OPCS4-M28.8 | Other specified |
| | OPCS4-M28.9 | Unspecified |
| | OPCS4- M31 | Extracorporeal fragmentation of calculus of ureter |
| | OPCS4-M31.1 | Extracorporeal shockwave lithotripsy of calculus of ureter |
| | OPCS4-M31.8 | Other specified |
| | OPCS4-M31.9 | Unspecified |
| Self-reported operation | 1197 | Percutaneous/open kidney stone surgery/lithotripsy |
| Death codes | N20.0 | Calculus of kidney |
| | N20.1 | Calculus of ureter |
| | N20.2 | Calculus of kidney with calculus of ureter |
| | N20.9 | Urinary calculus, unspecified |
| | N23 | Unspecified renal colic |
| Primary care Read V2 and V3 codes | Read V2-14D3. | H/O: urinary stone |
| | Read V2-1A54. | Ureteric colic |
| | Read V2-4G6.. | O/E - ureteric calculus |
| | Read V2-7B07. | Percutaneous renal stone surgery |
| | Read V2-7B070 | Nephroscopy and ultrasound lithotripsy of renal calculus |
| | Read V2-7B071 | Nephroscopy and electrohydraulic lithotripsy of renal calculus |
| | Read V2-7B072 | Nephroscopy and laser lithotripsy of renal calculus |
| | Read V2-7B073 | Percutaneous nephrolithotomy without disintegration |
| | Read V2-7B074 | Endoscopic extraction of calculus of kidney nec |
| | Read V2-7B07y | Other specified percutaneous renal stone surgery |
| | Read V2-7B07z | Percutaneous renal stone surgery NOS |

| | | |
|---|---|---|
| | Read V2-7B0B. | Extracorporeal shockwave lithotripsy for renal calculus |
| | Read V2-7B0B0 | Extracorporeal shockwave lithotripsy for renal calculus of unspecified size |
| | Read V2-7B0B1 | Extracorporeal shockwave lithotripsy for renal calculus less than 2 cm in diameter |
| | Read V2-7B0B2 | Extracorporeal shockwave treatment for renal calculus of 2 cm or more in diameter |
| | Read V2-7B0By | Other specified extracorporeal shockwave lithotripsy for renal calculus |
| | Read V2-7B0Bz | Extracorporeal shockwave lithotripsy for renal calculus NOS |
| | Read V2-7B170 | Nephroscopic laser lithotripsy of ureteric calculus |
| | Read V2-7B171 | Other nephroscopic fragmentation of ureteric calculus |
| | Read V2-7B172 | Nephroscopic extraction of ureteric calculus |
| | Read V2-7B18. | Ureteroscopic operations for ureteric calculus |
| | Read V2-7B180 | Ureteroscopic laser lithotripsy of ureteric calculus |
| | Read V2-7B181 | Other ureteroscopic fragmentation of ureteric calculus |
| | Read V2-7B182 | Ureteroscopic extraction of ureteric calculus |
| | Read V2-7B19. | Cystoscopic removal of ureteric calculus |
| | Read V2-7B190 | Cystoscopic laser lithotripsy of ureteric calculus |
| | Read V2-7B191 | Other cystoscopic fragmentation of ureteric calculus |
| | Read V2-7B193 | Cystoscopic catheter drainage for ureteric calculus |
| | Read V2-7B194 | Cystoscopic dilation of ureter for drainage of calculus |
| | Read V2-7B19y | Other specified cystoscopic removal of ureteric calculus |
| | Read V2-7B19z | Cystoscopic removal of ureteric calculus NOS |
| | Read V2-7B1C. | Extracorporeal shockwave lithotripsy of ureteric calculus |
| | Read V2-7B1C0 | Extracorporeal shockwave lithotripsy of unspecified ureteric calculus |

| | | |
|---|---|---|
| | Read V2-7B1C1 | Extracorporeal shockwave therapy for stone in upper ureter |
| | Read V2-7B1C2 | Extracorporeal shockwave lithotripsy for stone in mid-ureter |
| | Read V2-7B1C3 | Extracorporeal shockwave lithotripsy for stone in lower ureter |
| | Read V2-7B1Cy | Other specified extracorporeal shockwave lithotripsy of ureteric calculus |
| | Read V2-7B1Cz | Extracorporeal shockwave lithotripsy of ureteric calculus NOS |
| | Read V2-C3411 | Uric acid nephrolithiasis |
| | Read V2-K112. | Hydronephrosis with renal and ureteral calculous obstruction |
| | Read V2-K12.. | Calculus of kidney and ureter |
| | Read V2-K120. | Calculus of kidney |
| | Read V2-K1200 | Staghorn calculus |
| | Read V2-K120z | Renal calculus NOS |
| | Read V2-K121. | Calculus of ureter |
| | Read V2-K122. | Calculus of kidney with calculus of ureter |
| | Read V2-K12z. | Urinary calculus NOS |
| | Read V2-Kyu3. | [X]Urolithiasis |
| | Read V2-R080. | [D]Renal colic |
| | Read V2-R0800 | [D]Renal colic, unspecified |
| | Read V2-R0801 | [D]Ureteric colic |
| | Read V2-R080z | [D]Renal colic NOS |
| | Read V3-14D3. | H/O: urinary stone |
| | Read V3-4G6.. | O/E - ureteric calculus |
| | Read V3-7B07. | Percutaneous nephrolithotomy |

| | | |
|---|---|---|
| | Read V3-7B070 | Nephroscopy and ultrasound lithotripsy of renal calculus |
| | Read V3-7B071 | Nephroscopy and electrohydraulic lithotripsy of renal calculus |
| | Read V3-7B072 | Endoscopic laser fragmentation of renal calculus |
| | Read V3-7B073 | Percutaneous nephrolithotomy without disintegration |
| | Read V3-7B07y | Other specified percutaneous renal stone surgery |
| | Read V3-7B07z | Percutaneous renal stone surgery NOS |
| | Read V3-7B0B. | Extracorporeal shockwave lithotripsy for renal calculus |
| | Read V3-7B0B0 | Extracorporeal shockwave lithotripsy for renal calculus of unspecified size |
| | Read V3-7B0B1 | Extracorporeal shockwave lithotripsy for renal calculus less than 2 cm in diameter |
| | Read V3-7B0B2 | Extracorporeal shockwave treatment for renal calculus of 2 cm or more in diameter |
| | Read V3-7B0By | Other specified extracorporeal shockwave lithotripsy for renal calculus |
| | Read V3-7B0Bz | Extracorporeal shockwave lithotripsy for renal calculus NOS |
| | Read V3-7B170 | Nephroscopic laser fragmentation of ureteric calculus |
| | Read V3-7B171 | Other nephroscopic fragmentation of ureteric calculus |
| | Read V3-7B172 | Nephroscopic removal of ureteric calculus |
| | Read V3-7B18. | Ureteroscopic operations for ureteric calculus |
| | Read V3-7B180 | Ureteroscopic laser lithotripsy of ureteric calculus |
| | Read V3-7B181 | Other ureteroscopic fragmentation of ureteric calculus |
| | Read V3-7B182 | Ureteroscopic extraction of ureteric calculus |
| | Read V3-7B19. | Cystoscopic operation for ureteric calculus |
| | Read V3-7B190 | Cystoscopic laser lithotripsy of ureteric calculus |
| | Read V3-7B191 | Other cystoscopic fragmentation of ureteric calculus |

| | | |
|---|---|---|
| | Read V3-7B193 | Cystoscopic catheter drainage for ureteric calculus |
| | Read V3-7B194 | Cystoscopic dilation of ureter for drainage of calculus |
| | Read V3-7B19y | Other specified cystoscopic removal of ureteric calculus |
| | Read V3-7B19z | Cystoscopic removal of ureteric calculus NOS |
| | Read V3-7B1C. | Extracorporeal shockwave lithotripsy of ureteric calculus |
| | Read V3-7B1C0 | Extracorporeal shockwave lithotripsy of unspecified ureteric calculus |
| | Read V3-7B1C1 | Extracorporeal shockwave therapy for stone in upper ureter |
| | Read V3-7B1C2 | Extracorporeal shockwave lithotripsy for stone in mid-ureter |
| | Read V3-7B1C3 | Extracorporeal shockwave lithotripsy for stone in lower ureter |
| | Read V3-7B1Cy | Other specified extracorporeal shockwave lithotripsy of ureteric calculus |
| | Read V3-7B1Cz | Extracorporeal shockwave lithotripsy of ureteric calculus NOS |
| | Read V3-C3411 | Renal stone - uric acid |
| | Read V3-K112. | Hydronephrosis with renal and ureteral calculous obstruction |
| | Read V3-K1200 | Staghorn calculus |
| | Read V3-K120z | Renal calculus NOS |
| | Read V3-K121. | Calculus of ureter |
| | Read V3-K12z. | Urinary calculus NOS |
| | Read V3-R080. | [D]Renal colic |
| | Read V3-R0800 | [D]Renal colic, unspecified |
| | Read V3-R0801 | [D]Ureteric colic |
| | Read V3-R080z | [D]Renal colic NOS |
| | Read V3-X30Pl | Urolithiasis |

| | | |
|---|---|---|
| | Read V3-X30Pm | Urinary calculus |
| | Read V3-X30Pn | Nephrolithiasis NOS |
| | Read V3-Xa07P | C/O - ureteric pain |
| | Read V3-Xa6m6 | Ureteroscopic operation for ureteric calculus |
| | Read V3-Xa8P3 | Cystoscopic extraction of ureteric calculus without disintegration |
| | Read V3-XE0dj | Calculus of kidney and ureter |
| | Read V3-XE0dk | Kidney calculus |
| | Read V3-XE0G7 | Ureteroscopic removal of ureteric calculus |
| | Read V3-XE0G8 | Cystoscopic extraction of ureteric calculus |
| | Read V3-XE2Pu | Ureteric colic |
| | Read V3-XM0CQ | C/O - ureteric colic |

*Supplementary Table S2. Inheritance modes of 34 gene kidney stone panel R256 “Nephrocalcinosis or nephrolithiasis” (Version: 5.0)*

| **Gene** | **Gene Name** | **Inheritance** |
|---|---|---|
| *AGXT* | Alanine-Glyoxylate Aminotransferase | AR |
| *APRT* | Adenine Phosphoribosyltransferase | AR |
| *ATP6V0A4* | ATPase H+ Transporting V0 Subunit A4 | AR |
| *ATP6V1B1* | ATPase H+ Transporting V1 Subunit B1 | AR |
| *BSND* | Barttin CLCNK Type Accessory Subunit Beta | AR |
| *CA2* | Carbonic Anhydrase 2 | AR |
| *CASR* | Calcium-Sensing Receptor | AD |
| *CLCN5* | Chloride Voltage-Gated Channel 5 | XLR |
| *CLCNKB* | Chloride Voltage-Gated Channel Kb | AR |
| *CLDN16* | Claudin 16 | AR |
| *CLDN19* | Claudin 19 | AR |
| *CYP24A1* | Cytochrome P450 Family 24 Subfamily A Member 1 | AD/AR |
| *FAM20A* | FAM20A Golgi Associated Secretory Pathway Pseudokinase | AR |
| *GRHPR* | Glyoxylate And Hydroxypyruvate Reductase | AR |
| *HNF4A* | Hepatocyte Nuclear Factor 4 Alpha | AD* |
| *HOGA1* | 4-Hydroxy-2-Oxoglutarate Aldolase | AR |
| *HPRT1* | Hypoxanthine-Guanine Phosphoribosyltransferase 1 | XLR |
| *KCNJ1* | Potassium Inwardly Rectifying Channel Subfamily J Member 1 | AR |
| *MOCOS* | Molybdenum Cofactor Sulfurase | AR |
| *OCRL* | Inositol Polyphosphate 5-Phosphatase | XLR |
| *PHEX* | Phosphate Regulating Endopeptidase X-Linked | XLD |
| *RRAGD* | RAS Related GTP Binding D | AD |
| *SLC12A1* | Solute Carrier Family 12 Member 1 | AR |
| *SLC22A12* | Solute Carrier Family 22 Member 12 | AR |
| *SLC2A9* | Solute Carrier Family 2 Member 9 | AD/AR |
| *SLC34A1* | Solute Carrier Family 34 Member 1 | AD/AR |
| *SLC34A3* | Solute Carrier Family 34 Member 3 | AD/AR |
| *SLC3A1* | Solute Carrier Family 3 Member 1 | AD/AR |
| *SLC4A1* | Solute Carrier Family 4 Member 1 | AD |

| | | |
|---|---|---|
| *SLC7A9* | Solute Carrier Family 7 Member 9 | AD/AR |
| *STRADA* | STE20 Related Adaptor Alpha | AR |
| *VIPAS39* | VPS33B Interacting Protein, Apical-Basolateral Polarity Regulator, Spe-39 Homolog | AR |
| *VPS33B* | VPS33B Late Endosome And Lysosome Associated | AR |
| *WDR72*† | WD repeat domain 72 | AR |
| *XDH* | Xanthine Dehydrogenase | AR |

AD= autosomal dominant; AR= autosomal recessive; XLD= x-linked dominant; XLR= x-linked recessive. * Arg63Trp (R63W) variant only as there is no evidence to support other variants being linked to KSD. †*WDR72* not included in analyses of the Newcastle cohort. Underlined text indicates inheritance mode proposed based on the findings of this study.

*Supplementary Table S3. Baseline characteristics of kidney stone disease cases and controls in the UK Biobank*

| | Overall (N=469,295) | Case (N=13,681) | Control (N=455,614) | P |
|---|---|---|---|---|
| **Sex** | | | | |
| Female | 254399 (54.2%) | 4618 (33.8%) | 249781 (54.8%) | <0.001 |
| Male | 214896 (45.8%) | 9063 (66.2%) | 205833 (45.2%) | |
| **Genetic ethnic grouping** | | | | |
| Caucasian | 393822 (83.9%) | 11431 (83.6%) | 382391 (83.9%) | 0.24 |
| Other | 75473 (16.1%) | 2250 (16.4%) | 73223 (16.1%) | |
| **Age at recruitment (years)** | | | | |
| Mean (SD) | 56.5 (8.09) | 57.7 (7.81) | 56.5 (8.10) | <0.001 |
| **Age at first KSD code (years)** | | | | |
| Mean (SD) | | 61.0 (11.1) | NA | |
| **Albumin-adjusted serum calcium concentration (mmol/L)** | | | | |
| Mean (SD) | 2.28 (0.08) | 2.281 (0.09) | 2.276 (0.08) | <0.001 |
| Missing | 59585 (12.7%) | 1691 (12.4%) | 57894 (12.7%) | |
| **Serum phosphate concentration (mmol/L)** | | | | |
| Mean (SD) | 1.16 (0.161) | 1.12 (0.172) | 1.16 (0.161) | <0.001 |
| Missing | 60047 (12.8%) | 1704 (12.5%) | 58343 (12.8%) | |
| **Serum 25-OH vitamin D concentration (nmol/L)** | | | | |
| Mean (SD) | 48.6 (21.1) | 47.2 (20.6) | 48.7 (21.1) | <0.001 |
| Missing | 41808 (8.9%) | 1177 (8.6%) | 40631 (8.9%) | |
| **Serum urate (µmol/L)** | | | | |
| Mean (SD) | 309 (80.5) | 333 (81.8) | 309 (80.3) | <0.001 |
| Missing | 22123 (4.7%) | 660 (4.8%) | 21463 (4.7%) | |
| **Serum creatinine (µmol/L)** | | | | |
| Mean (SD) | 72.3 (18.6) | 76.7 (22.5) | 72.2 (18.5) | <0.001 |
| Missing | 21804 (4.6%) | 655 (4.8%) | 21149 (4.6%) | |
| **Body mass index (kg/m$^2$)** | | | | |
| Mean (SD) | 27.4 (4.77) | 28.8 (5.07) | 27.4 (4.76) | <0.001 |
| Missing | 1891 (0.4%) | 108 (0.8%) | 1783 (0.4%) | |
| **Waist-to-hip ratio** | | | | |

| | Overall (N=469,295) | Case (N=13,681) | Control (N=455,614) | P |
|---|---|---|---|---|
| Mean (SD) | 0.87 (0.09) | 0.91 (0.09) | 0.87 (0.09) | <0.001 |
| Missing | 1120 (0.2%) | 67 (0.5%) | 1053 (0.2%) | |

*Supplementary Table S4. Baseline characteristics of single and recurrent kidney stone disease cases in the UK Biobank*

| | Overall (N=14,238) | Non-recurrent stone former (N=9,475) | Recurrent stone former (N=4,763) | P |
|---|---|---|---|---|
| **Sex** | | | | |
| Female | 4819 (33.8%) | 3390 (35.8%) | 1429 (30.0%) | <0.001 |
| Male | 9419 (66.2%) | 6085 (64.2%) | 3334 (70.0%) | |
| **Genetic ethnic grouping** | | | | |
| Caucasian | 11898 (83.6%) | 7862 (83.0%) | 4036 (84.7%) | <0.001 |
| Other | 2340 (16.4%) | 1613 (17.0%) | 727 (15.3%) | |
| **Age at recruitment (years)** | | | | |
| Mean (SD) | 57.7 (7.79) | 57.6 (7.84) | 57.9 (7.69) | 0.070 |
| **Age at first KSD code (years)** | | | | |
| Mean (SD) | 61.1 (11.1) | 61.2 (11.3) | 60.9 (10.9) | 0.221 |
| **Albumin-adjusted serum calcium concentration (mmol/L)** | | | | |
| Mean (SD) | 2.28 (0.092) | 2.28 (0.091) | 2.28 (0.093) | 0.146 |
| Missing | 1777 (12.5%) | 1167 (12.3%) | 610 (12.8%) | |
| **Serum phosphate concentration (mmol/L)** | | | | |
| Mean (SD) | 1.12 (0.172) | 1.12 (0.172) | 1.11 (0.172) | <0.001 |
| Missing | 1791 (12.6%) | 1174 (12.4%) | 617 (13.0%) | |
| **Serum 25-OH vitamin D concentration (nmol/L)** | | | | |
| Mean (SD) | 47.2 (20.6) | 47.5 (20.6) | 46.6 (20.6) | 0.013 |
| Missing | 1234 (8.7%) | 862 (9.1%) | 372 (7.8%) | |
| **Serum urate (µmol/L)** | | | | |
| Mean (SD) | 333 (81.6) | 330 (82.0) | 337 (80.7) | <0.001 |
| Missing | 703 (4.9%) | 489 (5.2%) | 214 (4.5%) | |
| **Serum creatinine (µmol/L)** | | | | |
| Mean (SD) | 76.7 (22.5) | 76.4 (23.5) | 77.4 (20.3) | 0.012 |
| Missing | 697 (4.9%) | 488 (5.2%) | 209 (4.4%) | |
| **Body mass index (kg/m$^2$)** | | | | |
| Mean (SD) | 28.8 (5.10) | 28.6 (5.09) | 29.1 (5.12) | <0.001 |
| Missing | 112 (0.8%) | 69 (0.7%) | 43 (0.9%) | |

| | Overall (N=14,238) | Non-recurrent stone former (N=9,475) | Recurrent stone former (N=4,763) | P |
|---|---|---|---|---|
| **Waist-to-hip ratio** | | | | |
| Mean (SD) | 0.914 (0.0872) | 0.910 (0.089) | 0.923 (0.083) | <0.001 |
| Missing | 70 (0.5%) | 33 (0.3%) | 37 (0.8%) | |

*Supplementary Table S5. Distribution of variant pathogenicity in the  kidney stone disease gene panel R256 from UK Biobank*

| Gene | Number of pathogenic/likely pathogenic heterozygous variants identified in InterVar and extracted from UK Biobank population | | |
|---|---|---|---|
| | **Total** | **Likely Pathogenic** | **Pathogenic** |
| *AGXT* | 37 | 12 | 25 |
| *APRT* | 27 | 17 | 10 |
| *ATP6V0A4* | 50 | 9 | 41 |
| *ATP6V1B1* | 40 | 19 | 21 |
| *BSND* | 18 | 13 | 5 |
| *CA2* | 12 | 6 | 6 |
| *CASR* | 40 | 21 | 19 |
| *CLCNKB* | 66 | 26 | 40 |
| *CLDN16* | 11 | 4 | 7 |
| *CLDN19* | 18 | 10 | 8 |
| *CYP24A1* | 50 | 17 | 33 |
| *FAM20A* | 35 | 6 | 29 |
| *GRHPR* | 31 | 11 | 20 |
| *HNF4A* | 0 | 0 | 0 |
| *HOGA1* | 20 | 7 | 13 |
| *KCNJ1* | 25 | 11 | 14 |
| *MOCOS* | 56 | 14 | 42 |
| *SLC12A1* | 54 | 17 | 37 |
| *SLC22A12* | 127 | 101 | 26 |
| *SLC34A1* | 43 | 14 | 29 |
| *SLC34A3* | 49 | 17 | 32 |
| *SLC3A1* | 68 | 28 | 40 |
| *SLC4A1* | 28 | 10 | 18 |
| *SLC7A9* | 39 | 24 | 15 |
| *STRADA* | 17 | 4 | 13 |
| *VIPAS39* | 39 | 5 | 34 |
| *VPS33B* | 50 | 7 | 43 |

| | | | |
|---|---|---|---|
| *WDR72* | 65 | 18 | 47 |
| *XDH* | 92 | 29 | 63 |
| **Total** | **1207** | **477** | **730** |

X-linked genes CLCN5 and OCRL1 were excluded. No variants in HPRT1, PHEX. RRAGD or SLC2A9 were identified in UK Biobank genetic data.

***Supplementary Table S6. Proportion of Mendelian diagnoses of kidney stone disease in UK Biobank associated with CYP24A1, SLC2A9, SLC34A1, SLC34A3, SLC3A1, and SLC7A9 using two modes of inheritance***

| Model | Gene | Genetic diagnosis | All UK Biobank | Controls | Single KSD | Recurrent KSD | Total KSD cases | % with KSD | Trend | Single vs recurrent | Recurrent vs control | Single vs control | KSD vs control |
|---|---|---|---|---|---|---|---|---|---|---|---|---|---|
| Monoallelic inheritance | *CYP24A1* | Yes | 906 | 868 | 34 | 4 | 38 | 4.19% | 0.18 | 1.00 | 1.00 | 0.07 | 0.14 |
| | | No | 468389 | 454746 | 11390 | 2253 | 13643 | 2.91% | | | | | |
| | | % with diagnosis | 0.19% | 0.19% | 0.30% | 0.18% | 0.28% | | | | | | |
| | *SLC34A1* | Yes | 517 | 502 | 9 | 6 | 15 | 2.90% | 0.25 | 0.18 | 0.29 | 1.00 | 1.00 |
| | | No | 468778 | 455112 | 11415 | 2251 | 13666 | 2.92% | | | | | |
| | | % with diagnosis | 0.11% | 0.11% | 0.08% | 0.27% | 0.11% | | | | | | |
| | *SLC34A3* | Yes | 853 | 802 | 43 | 8 | 51 | 5.98% | 3.42x10-6 | 1.00 | 0.39 | 5.69x10-6 | 8.87x10-7 |
| | | No | 468442 | 454812 | 11381 | 2249 | 13630 | 2.91% | | | | | |
| | | % with diagnosis | 0.18% | 0.18% | 0.38% | 0.35% | 0.37% | | | | | | |
| | *SLC3A1* | Yes | 0 | 0 | 0 | 0 | 0 | *NA* | *NA* | *NA* | *NA* | *NA* | *NA* |
| | | No | 469295 | 455614 | 11424 | 2257 | 13681 | 2.92% | | | | | |
| | | % with diagnosis | 0.00% | 0.00% | 0.00% | 0.00% | 0.00% | | | | | | |
| | *SLC7A9* | Yes | 2795 | 2707 | 65 | 23 | 88 | 3.15% | 0.15 | 0.11 | 0.07 | 1.00 | 1.00 |
| | | No | 466500 | 452907 | 11359 | 2234 | 13593 | 2.91% | | | | | |
| | | % with diagnosis | 0.60% | 0.59% | 0.57% | 1.02% | 0.64% | | | | | | |
| Biallelic inheritance | *CYP24A1* | Yes | 1 | 1 | 0 | 0 | 0 | 0.00% | 0.11 | *NA* | 1.00 | 1.00 | 1.00 |
| | | No | 469294 | 455613 | 11424 | 2257 | 13681 | 2.92% | | | | | |
| | | % with diagnosis | 0.00% | 0.00% | 0.00% | 0.00% | 0.00% | | | | | | |
| | *SLC34A1* | Yes | 1 | 1 | 0 | 0 | 0 | 0.00% | 0.14 | *NA* | 1.00 | 1.00 | 1.00 |
| | | No | 469294 | 455613 | 11424 | 2257 | 13681 | 2.92% | | | | | |
| | | % with diagnosis | 0.00% | 0.00% | 0.00% | 0.00% | 0.00% | | | | | | |

| | | | | | | | | | | | | |
|---|---|---|---|---|---|---|---|---|---|---|---|---|
| *SLC34A3* | Yes | 0 | 0 | 0 | 0 | 0 | *NA* | *NA* | *NA* | *NA* | *NA* | *NA* |
| | No | 469295 | 455614 | 11424 | 2257 | 13681 | 2.92% | | | | | |
| | % with diagnosis | 0.00% | 0.00% | 0.00% | 0.00% | 0.00% | | | | | | |
| *SLC3A1* | Yes | 0 | 0 | 0 | 0 | 0 | *NA* | *NA* | *NA* | *NA* | *NA* | *NA* |
| | No | 469295 | 455614 | 11424 | 2257 | 13681 | 2.92% | | | | | |
| | % with diagnosis | 0.00% | 0.00% | 0.00% | 0.00% | 0.00% | | | | | | |
| *SLC7A9* | Yes | 4 | 2 | 1 | 1 | 2 | 50.00% | 0.00 | 1.00 | 0.07 | 0.36 | 0.02 |
| | No | 469291 | 455612 | 11423 | 2256 | 13679 | 2.91% | | | | | |
| | % with diagnosis | 0.00% | 0.00% | 0.01% | 0.04% | 0.01% | | | | | | |

No variants in SLC2A9 were identified in UK Biobank genetic data

***Supplemental Table S7. Genotype-phenotype correlations in individuals with P/LP variants in SLC34A1, SLC34A3, and CYP24A1 in the UK Biobank assuming monoallelic inheritance***

| Gene | | Genetic diagnosis (N=517) | No genetic diagnosis (N=468778) | P |
|---|---|---|---|---|
| ***SLC34A1*** | **Sex** | | | |
| | Female | 280 (54.2%) | 254119 (54.2%) | 1.00 |
| | Male | 237 (45.8%) | 214659 (45.8%) | |
| | **Age at first stone** | | | |
| | Mean (SD) | 61.7 (13.1) | 61.0 (11.1) | 0.87 |
| | Missing | 505 (97.7%) | 458790 (97.9%) | |
| | **KSD status** | | | |
| | Case | 15 (2.9%) | 13666 (2.9%) | 1.00 |
| | Control | 502 (97.1%) | 455112 (97.1%) | |
| | **Albumin-adjusted serum calcium concentration (mmol/L)** | | | |
| | Mean (SD) | 2.28 (0.0781) | 2.28 (0.0820) | 0.31 |
| | Missing | 73 (14.1%) | 59512 (12.7%) | |
| | **Serum phosphate concentration (mmol/L)** | | | |
| | Mean (SD) | 1.14 (0.161) | 1.16 (0.161) | **0.01** |
| | Missing | 73 (14.1%) | 59974 (12.8%) | |
| | | **Genetic diagnosis (N=853)** | **No genetic diagnosis (N=468442)** | **P** |
| ***SLC34A3*** | **Sex** | | | |
| | Female | 478 (56.0%) | 253921 (54.2%) | 0.30 |
| | Male | 375 (44.0%) | 214521 (45.8%) | |
| | **Age at first stone** | | | |
| | Mean (SD) | 63.8 (11.0) | 61.0 (11.1) | 0.13 |
| | Missing | 814 (95.4%) | 458481 (97.9%) | |
| | **KSD status** | | | |
| | Case | 51 (6.0%) | 13630 (2.9%) | **1.77x10-7** |
| | Control | 802 (94.0%) | 454812 (97.1%) | |
| | **Albumin-adjusted serum calcium concentration (mmol/L)** | | | |
| | Mean (SD) | 2.28 (0.0808) | 2.28 (0.0820) | 0.16 |
| | Missing | 109 (12.8%) | 59476 (12.7%) | |
| | **Serum phosphate concentration (mmol/L)** | | | |
| | Mean (SD) | 1.11 (0.157) | 1.16 (0.161) | **7.83x10-18** |
| | Missing | 114 (13.4%) | 59933 (12.8%) | |
| | | **Genetic diagnosis (N=906)** | **No genetic diagnosis (N=468389)** | **P** |
| ***CYP24A1*** | **Sex** | | | |
| | Female | 511 (56.4%) | 253888 (54.2%) | 0.20 |
| | Male | 395 (43.6%) | 214501 (45.8%) | |
| | **Age at first stone** | | | |
| | Mean (SD) | 63.2 (10.2) | 61.0 (11.1) | 0.28 |
| | Missing | 878 (96.9%) | 458417 (97.9%) | |
| | **KSD status** | | | |
| | Case | 38 (4.2%) | 13643 (2.9%) | **0.03** |
| | Control | 868 (95.8%) | 454746 (97.1%) | |
| | **Albumin-adjusted serum calcium concentration (mmol/L)** | | | |
| | Mean (SD) | 2.29 (0.0911) | 2.28 (0.0820) | **2.67x10-4** |
| | Missing | 104 (11.5%) | 59481 (12.7%) | |
| | **Serum phosphate concentration (mmol/L)** | | | |
| | Mean (SD) | 1.17 (0.161) | 1.16 (0.161) | 0.06 |
| | Missing | 105 (11.6%) | 59942 (12.8%) | |

**Supplementary Table S8. Genotype-phenotype correlations in individuals with kidney stone disease and P/LP variants in SLC34A1, SLC34A3, and CYP24A1 in the UK Biobank assuming monoallelic inheritance**

| | | Genetic diagnosis (N=15) | No genetic diagnosis (N=13666) | P |
|---|---|---|---|---|
| *SLC34A1* | **Sex** | | | |
| | Female | 6 (40.0%) | 4612 (33.7%) | 0.81 |
| | Male | 9 (60.0%) | 9054 (66.3%) | |
| | **Age at first stone** | | | |
| | Mean (SD) | 61.7 (13.1) | 61.0 (11.1) | 0.87 |
| | Missing | 3 (20.0%) | 3678 (26.9%) | |
| | **KSD status** | | | |
| | Single episode KSD | 9 (60.0%) | 11415 (83.5%) | 0.04 |
| | Recurrent KSD | 6 (40.0%) | 2251 (16.5%) | |
| | **Albumin-adjusted serum calcium concentration (mmol/L)** | | | |
| | Mean (SD) | 2.33 (0.0824) | 2.28 (0.0915) | 0.07 |
| | Missing | 1 (6.7%) | 1690 (12.4%) | |
| | **Serum phosphate concentration (mmol/L)** | | | |
| | Mean (SD) | 1.03 (0.134) | 1.12 (0.172) | **0.03** |
| | Missing | 1.00 (6.7%) | 1703 (12.5%) | |
| | | **Genetic diagnosis (N=51)** | **No genetic diagnosis (N=13630)** | **P** |
| *SLC34A3* | **Sex** | | | |
| | Female | 16 (31.4%) | 4602 (33.8%) | 0.83 |
| | Male | 35 (68.6%) | 9028 (66.2%) | |
| | **Age at first stone** | | | |
| | Mean (SD) | 63.8 (11.0) | 61.0 (11.1) | 0.13 |
| | Missing | 12 (23.5%) | 3669 (26.9%) | |
| | **KSD status** | | | |
| | Single episode KSD | 43 (84.3%) | 11381 (83.5%) | 1.00 |
| | Recurrent KSD | 8 (15.7%) | 2249 (16.5%) | |
| | **Albumin-adjusted serum calcium concentration (mmol/L)** | | | |
| | Mean (SD) | 2.29 (0.0910) | 2.28 (0.0915) | 0.53 |
| | Missing | 4 (7.8%) | 1687 (12.4%) | |
| | **Serum phosphate concentration (mmol/L)** | | | |
| | Mean (SD) | 1.06 (0.167) | 1.12 (0.172) | **0.02** |
| | Missing | 4 (7.8%) | 1700 (12.5%) | |
| | | **Genetic diagnosis (N=38)** | **No genetic diagnosis (N=13643)** | **P** |
| *CYP24A1* | **Sex** | | | |
| | Female | 9 (23.7%) | 4609 (33.8%) | 0.25 |
| | Male | 29 (76.3%) | 9034 (66.2%) | |
| | **Age at first stone** | | | |
| | Mean (SD) | 63.2 (10.2) | 61.0 (11.1) | 0.28 |
| | Missing | 10 (26.3%) | 3671 (26.9%) | |
| | **KSD status** | | | |
| | Single episode KSD | 34 (89.5%) | 11390 (83.5%) | 0.44 |
| | Recurrent KSD | 4 (10.5%) | 2253 (16.5%) | |
| | **Albumin-adjusted serum calcium concentration (mmol/L)** | | | |
| | Mean (SD) | 2.26 (0.0710) | 2.28 (0.0916) | 0.14 |
| | Missing | 3 (7.9%) | 1688 (12.4%) | |
| | **Serum phosphate concentration (mmol/L)** | | | |
| | Mean (SD) | 1.16 (0.152) | 1.12 (0.172) | 0.11 |
| | Missing | 3 (7.9%) | 1701 (12.5%) | |

**Supplementary Table S9. Association of SLC34A1 variants with serum phosphate in UK Biobank assuming monoallelic inheritance**

| UK Biobank (Full cohort) | | | | |
|---|---|---|---|---|
| **Variable** | **Model** | **Estimate** | **SE** | **P** |
| **Carrier status** | Unadjusted | -0.02 | 0.01 | 0.01 |
| | Adjusted | -0.02 | 7.37x10-3 | 1.41x10-3 |
| **Age at recruitment** | Adjusted | 0.00 | 3.04x10-5 | 1.13x10-45 |
| **Creatinine** | Adjusted | 0.00 | 1.49x10-5 | 2.99x10-187 |
| **Adjusted calcium** | Adjusted | 0.23 | 3.03x10-3 | <2.20x10-16 |
| **Male sex** | Adjusted | -0.08 | 5.60x10-4 | <2.20x10-16 |
| **Kidney stone formers** | | | | |
| **Variable** | **Model** | **Estimate** | **SE** | **P** |
| **Carrier status** | Unadjusted | -0.09 | 0.05 | 0.05 |
| | Adjusted | -0.11 | 0.04 | 0.01 |
| **Age at recruitment** | Adjusted | 0.00 | 1.97x10-4 | 0.05 |
| **Creatinine** | Adjusted | 0.00 | 7.38x10-5 | 5.04x10-23 |
| **Adjusted calcium** | Adjusted | 0.11 | 0.02 | 8.88x10-10 |
| **Male sex** | Adjusted | -0.09 | 3.57x10-3 | 3.93x10-126 |

**Supplementary Table S10. Association of SLC34A3 variants with serum phosphate in UK Biobank assuming monoallelic inheritance**

| UK Biobank (Full cohort) | | | | |
|---|---|---|---|---|
| **Variable** | **Model** | **Estimate** | **SE** | **P** |
| **Carrier status** | Unadjusted | -0.05 | 0.01 | 1.18x10-17 |
| | Adjusted | -0.05 | 5.71x10-3 | 8.85x10-22 |
| **Age at recruitment** | Adjusted | 0.00 | 3.04x10-5 | 1.47x10-45 |
| **Creatinine** | Adjusted | 0.00 | 1.49x10-5 | 3.18x10-188 |
| **Adjusted calcium** | Adjusted | 0.23 | 3.03x10-3 | <2.20x10-16 |
| **Male sex** | Adjusted | -0.08 | 5.60x10-4 | <2.20x10-16 |
| **Kidney stone formers** | | | | |
| **Variable** | **Model** | **Estimate** | **SE** | **P** |
| **Carrier status** | Unadjusted | -0.06 | 0.03 | 0.02 |
| | Adjusted | -0.06 | 0.02 | 0.01 |
| **Age at recruitment** | Adjusted | 0.00 | 1.97x10-4 | 0.05 |
| **Creatinine** | Adjusted | 0.00 | 7.38x10-5 | 5.57x10-23 |
| **Adjusted calcium** | Adjusted | 0.11 | 0.02 | 1.02x10-9 |
| **Male sex** | Adjusted | -0.09 | 3.57x10-3 | 6.06x10-126 |

**Supplementary Table S11. Association of CYP24A1 variants with albumin-adjusted serum calcium in the UK Biobank assuming monoallelic inheritance**

| UK Biobank (Full cohort) | | | | |
|---|---|---|---|---|
| **Variable** | **Model** | **Estimate** | **SE** | **P** |
| **Carrier status** | Unadjusted | 0.01 | 0.01 | 0.06 |
| | Adjusted | 0.01 | 5.50x10-3 | 0.22 |
| **Age at recruitment** | Adjusted | 0.00 | 3.04x10-5 | 1.17x10-45 |
| **Creatinine** | Adjusted | 0.00 | 1.49x10-5 | 6.12x10-187 |
| **Phosphate** | Adjusted | 0.23 | 3.04x10-3 | <2.20x10-16 |
| **Male sex** | Adjusted | -0.08 | 5.60x10-4 | <2.20x10-16 |
| **Kidney stone formers** | | | | |
| **Variable** | **Model** | **Estimate** | **SE** | **P** |
| **Carrier status** | Unadjusted | -0.02 | 0.02 | 0.25 |
| | Adjusted | -0.02 | 0.02 | 0.22 |
| **Age at recruitment** | Adjusted | 0.00 | 1.04x10-4 | 3.57x10-38 |
| **Creatinine** | Adjusted | 0.00 | 3.92x10-5 | 1.64x10-12 |
| **Phosphate** | Adjusted | 0.03 | 4.85x10-3 | 9.80x10-10 |
| **Male sex** | Adjusted | -0.04 | 1.91x10-3 | 2.29x10-91 |

**Supplementary Table S12. Gene distribution of pathogenic/likely pathogenic variants identified from the R256 Nephrocalcinosis or Nephrolithiasis panel in individuals in the UK Biobank**

| Gene | Number of variants causing genetic diagnosis | |
|---|---|---|
| | UK Biobank | Individuals with kidney stone disease |
| *AGXT* | 2 | 0 |
| *ATP6V1B1* | 2 | 0 |
| *CASR* | 40 | 1 |
| *CLCNKB* | 1 | 0 |
| *CYP24A1* | 1 | 0 |
| *SLC22A12* | 6 | 0 |
| *SLC34A1* | 2 | 0 |
| *SLC34A3* | 49 | 11 |
| *SLC4A1* | 28 | 2 |
| *SLC7A9* | 39 | 10 |
| *WDR72* | 3 | 0 |
| *XDH* | 6 | 2 |

**Supplementary Table S13. P/LP Variants from the R256 Nephrocalcinosis or Nephrolithiasis panel accounting for a monogenic form of kidney stone disease identified in individuals with kidney stone disease in the UK Biobank**

| Gene | Variant | Genetic diagnosis | No genetic diagnosis | Missing genotype |
|---|---|---|---|---|
| *SLC7A9* | 19_32862207_C_CT | 33 | 13648 | 0 |
| *SLC34A3* | 9_137232928_G_A | 31 | 13650 | 0 |
| *SLC7A9* | 19_32864261_C_T | 19 | 13662 | 0 |
| *SLC7A9* | 19_32864206_G_A | 10 | 13671 | 0 |
| *SLC7A9* | 19_32843932_G_A | 9 | 13670 | 2 |
| *SLC7A9* | 19_32862151_G_A | 6 | 13674 | 1 |
| *SLC7A9* | 19_32859932_G_A | 5 | 13676 | 0 |
| *SLC34A3* | 9_137236173_T_TC | 5 | 13676 | 0 |
| *SLC7A9* | 19_32862554_G_A | 3 | 13678 | 0 |
| *SLC34A3* | 9_137232705_T_C | 3 | 13677 | 1 |
| *SLC34A3* | 9_137233358_A_AC | 3 | 13678 | 0 |
| *SLC7A9* | 19_32830686_T_C | 2 | 13678 | 1 |
| *SLC34A3* | 9_137232651_C_A | 2 | 13679 | 0 |
| *SLC34A3* | 9_137234733_T_A | 2 | 13679 | 0 |
| *SLC4A1* | 17_44254617_G_A | 1 | 13680 | 0 |
| *SLC4A1* | 17_44258596_G_A | 1 | 13680 | 0 |
| *SLC7A9* | 19_32830685_C_G | 1 | 13680 | 0 |
| *SLC7A9* | 19_32868486_G_A | 1 | 13680 | 0 |
| *XDH* | 2_31368641_A_ACC | 1 | 13680 | 0 |
| *XDH* | 2_31368643_A_AG | 1 | 13680 | 0 |
| *CASR* | 3_122284611_G_A | 1 | 13680 | 0 |
| *SLC34A3* | 9_137232783_G_A | 1 | 13680 | 0 |
| *SLC34A3* | 9_137234241_G_A | 1 | 13680 | 0 |
| *SLC34A3* | 9_137236219_C_T | 1 | 13680 | 0 |
| *SLC34A3* | 9_137236239_G_A | 1 | 13680 | 0 |
| *SLC34A3* | 9_137236380_C_G | 1 | 13680 | 0 |

**Supplemental Table S14. Kidney stone population attributable risk and population attributable fraction for pathogenic and likely pathogenic variants in the R256 Nephrocalcinosis or Nephrolithiasis panel in the UK Biobank**

| Gene | | Genetic diagnosis | No genetic diagnosis | Unexposed incidence | Exposed incidence | Total incidence | Population attributable risk | Population attributable fraction (%) |
|---|---|---|---|---|---|---|---|---|
| CASR | Controls | 99 | 455515 | 0.03 | 0.01 | 0.029 | -4.08x10-6 | -0.01 |
| | Cases | 1 | 13680 | | | | | |
| *SLC34A3* | Controls | 802 | 454812 | 0.03 | 0.04 | 0.029 | -0.97 | 0.19 |
| | Cases | 51 | 13630 | | | | | |
| *SLC4A1* | Controls | 50 | 455564 | 0.03 | 0.03 | 0.029 | 1.03x10-6 | 3.54x10-3 |
| | Cases | 2 | 13679 | | | | | |
| *SLC7A9* | Controls | 2707 | 452907 | 0.03 | 0.17 | 0.029 | 1.40x10-5 | 0.05 |
| | Cases | 88 | 13593 | | | | | |
| *XDH* | Controls | 5 | 455609 | 0.03 | 0.04 | 0.029 | 1.76x10-6 | 0.01 |
| | Cases | 1 | 13680 | | | | | |
| All | Controls | 3672 | 451942 | 0.03 | 0.04 | 0.029 | 6.83x10-5 | 0.23 |
| | Cases | 143 | 13538 | | | | | |

**Supplemental Table S15. Biochemical phenotype of individuals with kidney stone disease and an SLC34A3 diagnosis**

| | | Albumin adjusted serum calcium concentration | | |
|---|---|---|---|---|
| | | Low (<2.20 mmol/L) | Normal (2.20-2.60 mmol/L) | High (>2.60 mmol/L) |
| Serum phosphate concentration | Low (<0.80 mmol/L) | 0 | 4 | 0 |
| | Normal (0.80-1.50 mmol/L) | 4 | 39 | 0 |
| | High (>1.50 mmol/L) | 0 | 0 | 0 |

### Supplementary Figures

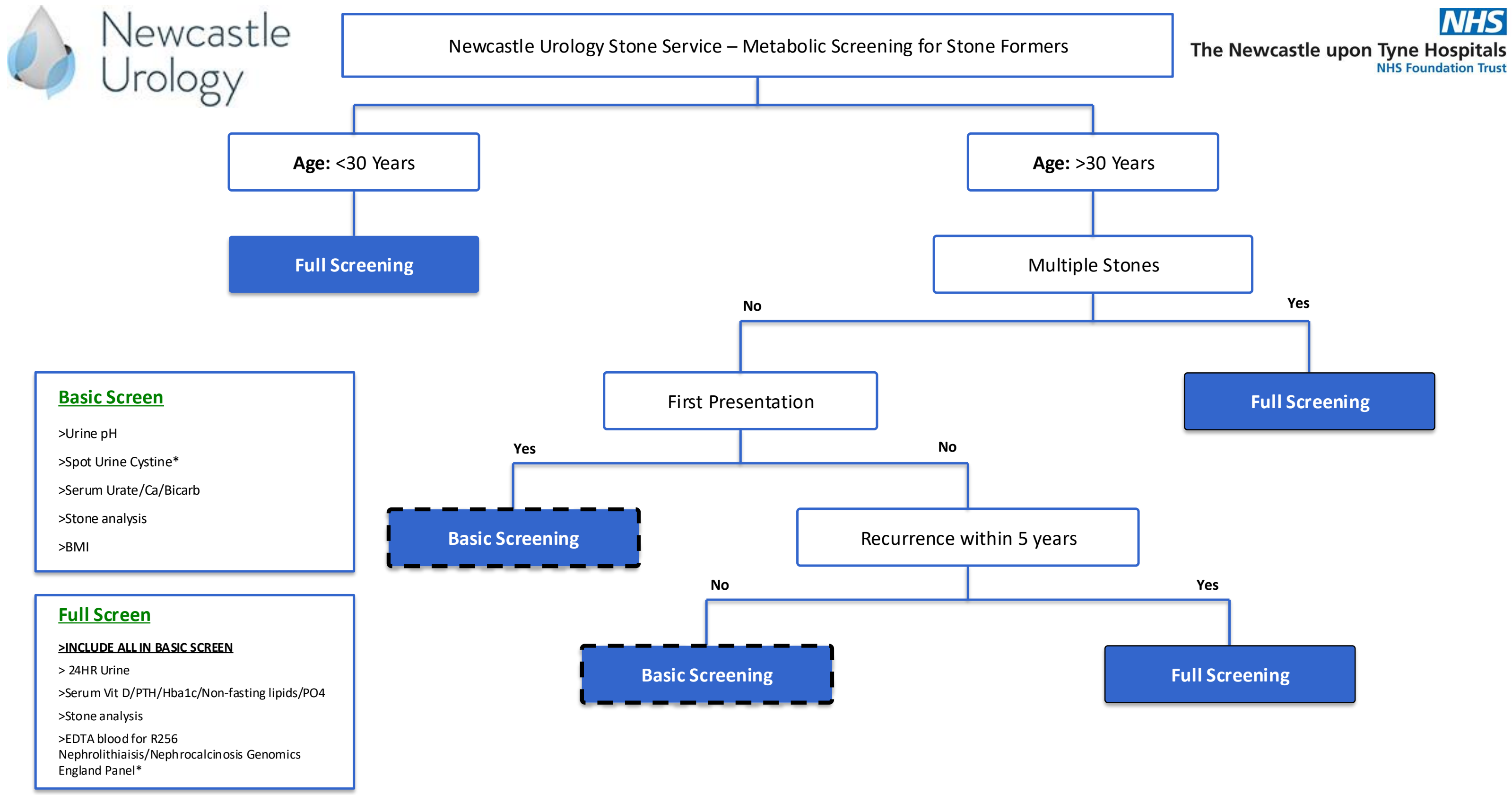

**Figure S1. Local referral pathway and work up of kidney stone disease patients via the Newcastle Urology Stone Service. All patients deemed to require 'Full Screening' were seen in the specialist nephrology clinic. Ca, calcium; Bicarb, bicarbonate; BMI, body mass index; Vit D, 25 hydroxyvitamin D; PTH, parathyroid hormone; PO4, phosphate.**

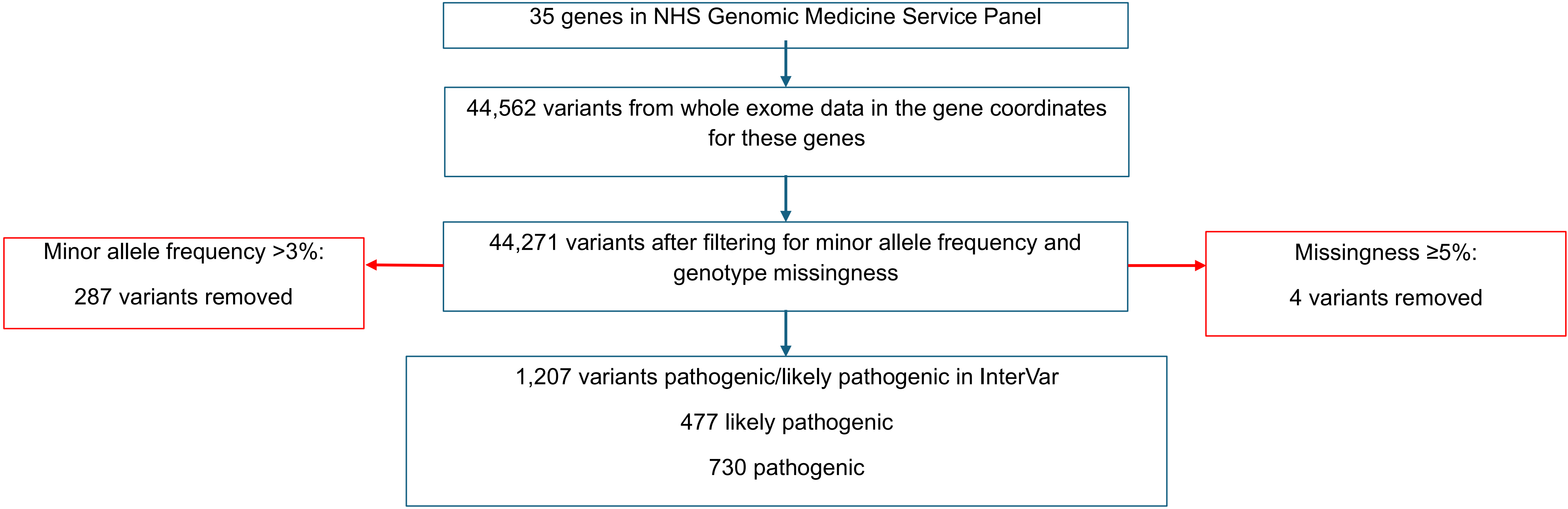

**Figure S2. Variant triage of alleles in 35 monogenic kidney stone disease genes using UK Biobank whole exome data**
